# Do drugs proposed to protect or harm joints change serious osteoarthritis events? A meta-analysis of adverse event tables from 881 randomized trials

**DOI:** 10.64898/2026.10.13.26365377

**Authors:** Zhida Xu, Mengjie Pan, Junhua Cheng, Xiaping Chen, Jie Luo, Xingrong Guo, Xiaohu Zhu

**Author notes:** **Correspondence to:** Dr Xingrong Guo,; Dr Xiaohu Zhu.

## Abstract

**Objectives:** Observational studies report large, conflicting associations between common drugs and hip or knee replacement. We tested them in the serious adverse event (SAE) tables of randomized trials on ClinicalTrials.gov.

**Methods:** We included randomized drug trials with ≥500 participants. A language model classified arms from summaries without outcome counts (masked by instruction only). We pooled osteoarthritis-related SAEs by drug class as Mantel–Haenszel risk ratios (RRs). Because the plan followed a preliminary analysis of 72 trials, we report estimates rather than tests, checked by positive and negative controls and calibration.

**Results:** The 881 placebo- or usual-care-controlled trials included 1.75 million participants and 9,408 osteoarthritis-related SAEs. The tables reproduced CANTOS’s own canakinumab effect (RR 0.57, 95% CI 0.39 to 0.82), the anti-nerve growth factor excess (2.31, 1.82 to 2.92) and two of three modest effects specified before extraction: gallbladder disease with glucagon-like peptide-1 receptor agonists (GLP-1RAs; 1.36, 1.22 to 1.51) and ketoacidosis with sodium– glucose cotransporter-2 (SGLT2) inhibitors (1.96, 1.48 to 2.60). In the same trials, osteoarthritis-related SAEs did not differ detectably with GLP-1RAs (0.97, 0.86 to 1.10; calibrated 0.87, 0.54 to 1.41) or SGLT2 inhibitors (0.96, 0.82 to 1.12; calibrated 0.86, 0.53 to 1.40). Other proposed drugs gave imprecise estimates. Post hoc, 8 of 21 published observational estimates differed from randomized ones, in both directions (3 to 6 after calibration).

**Conclusions:** Over about two years, GLP-1RAs and SGLT2 inhibitors had no detectable effect on osteoarthritis-related SAEs in trials that detected their known effects. Slow or modest structural effects remain possible.

**Key messages:** **What is already known on this topic**

- Observational studies associate glucose-lowering drugs, bisphosphonates and warfarin with large and conflicting changes in the risk of hip or knee replacement; randomized evidence is limited to a few exploratory analyses of single trials.

**What this study adds**

- GLP-1 receptor agonists and SGLT2 inhibitors had no detectable effect on serious osteoarthritis events over about two years, in randomized trials whose adverse-event tables detected these drugs’ known effects.

**How this study might affect research, practice or policy**

- Trial registries can test osteoarthritis repurposing signals for large, short-term effects before dedicated trials are funded; slow structural effects still need such trials.

## Introduction

Osteoarthritis affected 595 million people in 2020, and the number with knee or hip osteoarthritis is projected to rise by about three quarters by 2050.[1] Joint replacement, the treatment for end-stage disease, is common and costly, and no drug is approved to prevent it. Many people with osteoarthritis already take drugs for diabetes, obesity or cardiovascular disease, so finding one of these drugs that also protects the joint would be an attractive shortcut to disease modification. [2]

The evidence that such drugs exist comes mostly from routine-care data, and it points in many directions. In one health-record network, people with obesity and osteoarthritis who were prescribed GLP-1 receptor agonists (GLP-1RAs) had fewer hip and knee replacements, while those without osteoarthritis had more knee replacements.[3] Another analysis in the same network found more osteoarthritis among users with diabetes.[4] Active-comparator cohorts placed sodium–glucose cotransporter-2 (SGLT2) inhibitors below GLP-1RAs for joint replacement,[5, 6] and a nationwide cohort associated dipeptidyl peptidase-4 (DPP-4) inhibitors with 58–62% fewer replacements.[7] Warfarin was associated with more joint replacements than direct oral anticoagulants (DOACs),[8] and vitamin-K antagonist use with more radiographic osteoarthritis;[9] one proposed explanation is inhibition of vitamin-K-dependent matrix Gla protein. Bisphosphonates and statins have been associated with fewer knee replacements or with no difference.[10–12] All these associations are open to confounding by indication and to differential access to surgery: body mass index (BMI) causes osteoarthritis[13] and, where surgeons apply BMI thresholds, also determines who is eligible for arthroplasty,[14] so a drug that changes weight changes who is operated on.

Randomized evidence is scarce. Secondary analyses of single trials found fewer hip and knee replacements with canakinumab (CANTOS) and low-dose colchicine,[15, 16] more knee replacements during two years of treatment with zoledronic acid,[17] and no overall reduction with intensive lifestyle weight loss.[18] Anti-nerve growth factor (NGF) antibodies caused structural joint harm.[19, 20] Meta-analyses of osteoarthritis adverse events in GLP-1RA trials found no association,[21, 22] but whether such data can detect a true effect has not been tested.

A larger randomized record exists. Since the US Food and Drug Administration Amendments Act and its Final Rule, applicable trials must post serious adverse events (SAEs) by arm on ClinicalTrials.gov,[23] and these postings are more complete than journal reports.[24] Large trials of cardiometabolic drugs follow hundreds of thousands of older adults for years, and an elective joint replacement is a hospital admission that can appear among their SAEs. Registry SAE tables have been mined for repositioning signals before,[25] but, to our knowledge, not for osteoarthritis, and without systematic validation against known effects across drug classes. We therefore asked three questions in turn (Figure 1A). First, do these tables detect drug effects that are already known? Second, what do they show for the drugs proposed to protect or harm joints? Third, how do published observational estimates compare with the randomized ones?

**Figure 1.**
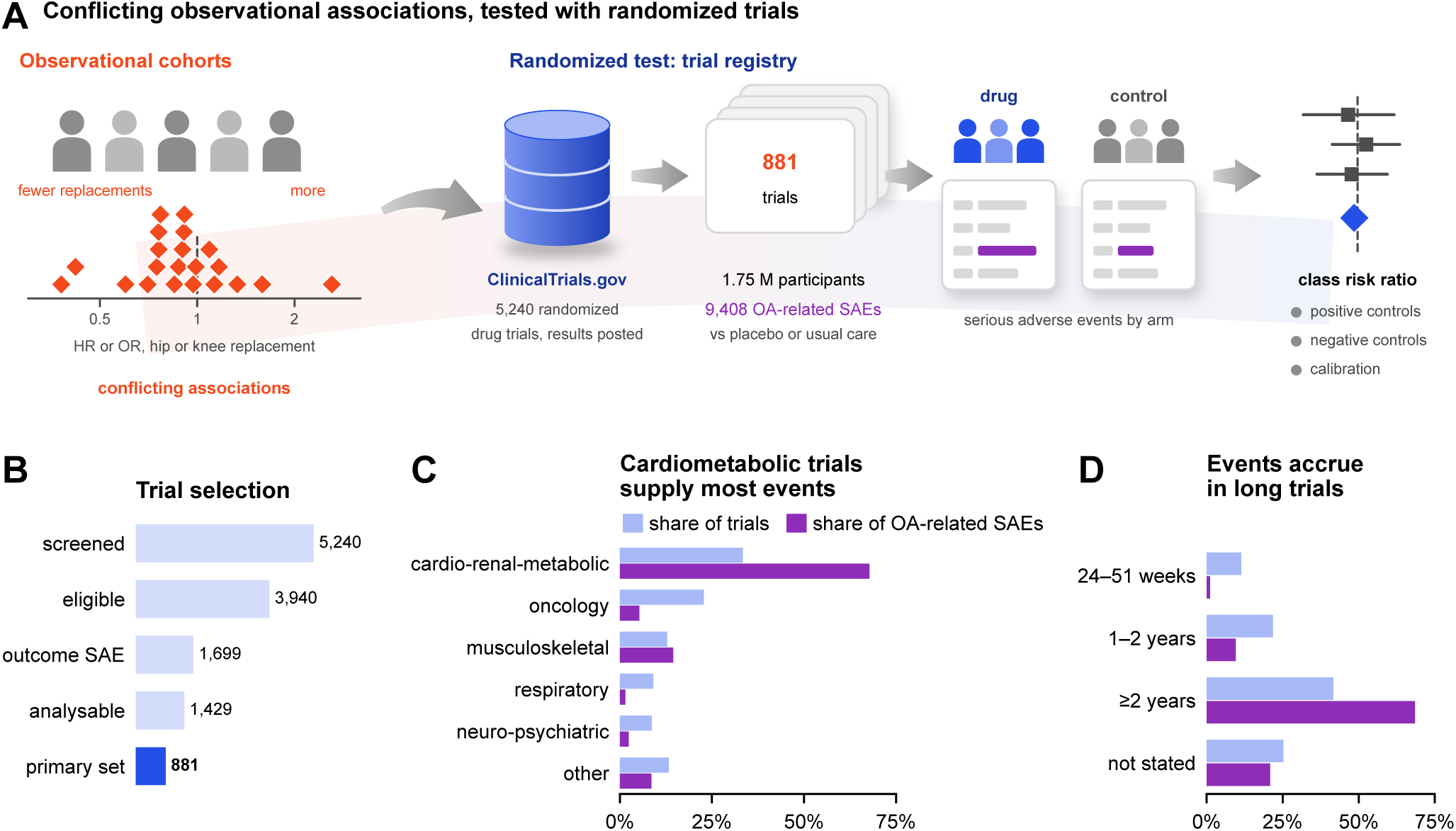
The question and the data. (A) Observational studies report large and conflicting associations between common drugs and hip or knee replacement (diamonds: the published estimates compared in Figure 5). We tested the same drug classes in the serious adverse event (SAE) tables of randomized trials posted on ClinicalTrials.gov: SAEs were counted by arm, pooled by drug class as Mantel–Haenszel risk ratios against placebo or usual care, and checked with positive controls, negative-control outcomes and empirical calibration (Figure 2). (B) Trial selection: randomized drug trials with at least 500 participants and posted adverse-event results (5,240), eligible (3,940), with an SAE of a protocol outcome in any arm and therefore adjudicated (1,699), contributing at least one randomized comparison (1,429), and placebo- or usual-care-controlled trials in the primary analysis (881; 1,751,625 participants; 9,408 osteoarthritis-related SAEs). The full flow is shown in Supplementary Figure S1. (C, D) Share of primary-analysis trials and of osteoarthritis-related SAEs by therapeutic area (C) and by stated adverse-event window (D). OA, osteoarthritis.

## Methods

### Protocol and its history

The analysis plan (Supplementary File 1) was time-stamped in a version-controlled repository on 26 September 2026 and was not registered externally. It was written after a preliminary pooled analysis of the SAE tables of 72 large trials, done while candidate questions were being screened (Supplementary File 3, with an English translation). That analysis already gave an estimate for one positive control (canakinumab) and for most hypotheses: those 72 trials contribute 87–98% of the events in the canakinumab and gout controls and in hypotheses H1– H3 and H8 (defined below). We therefore interpret the hypothesis tests as estimation, also report each estimate for the trials outside the preliminary analysis (Supplementary Table S7), and list every deviation (Supplementary Methods S7). Simulated internal and peer review and an automated re-audit, all by language models, prompted four rounds of error correction (Supplementary Table S5) and additional analyses; the latter were defined before they were run in three time-stamped addenda, referred to below as the first, second and third addenda (Supplementary Files 4, 6 and 7).

### Data source and eligibility

We used ClinicalTrials.gov (API version 2; records downloaded on 5 October 2026 and restricted to trials whose results were first posted by 26 September 2026, the date of the original query).[26] We included interventional, randomized trials with an enrolment of at least 500, posted adverse-event results and at least one drug or biological intervention. The size threshold favours trials with long follow-up and SAE surveillance, but it excludes smaller trials in osteoarthritis populations, such as STEP 9 (n = 407).[27] We excluded crossover, sequential and single-group designs, paediatric trials (minimum age below 16 years), vaccine trials and trials in orthopaedic or fracture-surgery populations. Trials with no SAE of the four protocol outcomes (osteoarthritis-related SAE, gout, cataract, inguinal hernia) in any arm carry no information for Mantel–Haenszel estimates and were not adjudicated.

### Arm adjudication

Adverse-event reporting groups were classified into randomized comparisons under written rules (Supplementary File 2): only core-period randomized groups were kept; doses of the same drug were pooled; add-on designs were analysed as placebo-controlled comparisons of the added drug, factorial trials by their margins, and active-comparator trials as head-to-head comparisons. A large language model (Claude Opus 5.5; Anthropic) did the adjudication on 26 September 2026 in eight parallel batches. It started from a rule-based pre-classification, worked from a structured summary of arms, interventions and reporting groups that did not show outcome counts, and recorded the drug under test and a confidence level. Masking was by instruction only: outcome tables were accessible in the model’s working environment, and the model recognized some trials by name. Structural checks confirmed that every trial was adjudicated once, with valid and non-overlapping groups. After an automated re-audit found misapplied rules, every included trial was screened for the same errors (third addendum; Supplementary Table S5): records that were sub-studies or extensions of other trials were excluded, and comparisons were dropped when a compared group included participants randomized to another arm or when the groups were reported over different periods by design (for example, placebo to week 16 against the drug to week 52). Two authors (ZX and MP) jointly checked a stratified sample of 202 trials against the registry records without outcome counts: every trial with at least 5 events in a positive-control or hypothesis analysis, the 20 primary-analysis trials with the most osteoarthritis-related SAEs, 100 randomly selected included trials and 20 randomly selected excluded trials. They confirmed the inclusion decision, group assignment and drug of all 202 trials, as corrected under the third addendum (100%; 95% CI 98.1 to 100).

### Drug classes and outcomes

Drugs were mapped to pre-specified mechanistic classes or, otherwise, to the fourth level of the WHO Anatomical Therapeutic Chemical classification (RxNav). The primary outcome, an osteoarthritis-related SAE, was any SAE term denoting osteoarthritis, osteoarthrosis, arthroplasty or joint replacement, including rapidly progressive osteoarthritis and chondrolysis, and excluding spinal, infective, prosthetic, fracture and traumatic terms. Terms for hand, foot and unspecified joints were included (Supplementary Methods S4).

This outcome is only a proxy for joint replacement. SAEs follow the E2A definition of the International Council for Harmonisation,[28] although some protocols exempt admissions planned before enrolment. Whether an elective replacement is reported therefore depends on the sponsor, and a replacement is often coded under the condition that led to it. The outcome is best read as admissions coded as osteoarthritis, most of them probably for replacement. We estimated capture (how completely the outcome was recorded) as control-arm events per 1,000 person-years, with the adverse-event window as follow-up. In CANTOS, we compared the recorded events with the replacements expected from the published trial. Counts were summed across matching terms, which can count a participant more than once; counting only each group’s most frequent term (maximum-term counting) gives a lower bound on the number of participants affected. Gout SAEs were the outcome of one positive control. Cataract and inguinal-hernia SAEs, elective surgical admissions in older adults, served as negative-control outcomes,[29] chosen on the assumption that none of the pre-specified drug classes affects them.

### Validation

Before interpreting any null result, we tested whether the tables recover known drug effects. The pre-specified positive controls were canakinumab (fewer osteoarthritis-related SAEs),[15] anti-NGF antibodies (more)[19] and SGLT2 inhibitors (fewer gout SAEs, expected from their reported reduction in gout outcomes).[30] The canakinumab control is not independent, because CANTOS ascertained replacements from the same trial database[15] and supplies 98% of the events. All three are large effects. The first addendum, written after the main results were known, therefore added five modest-effect controls on other SAEs; their expected sizes were taken from meta-analyses of randomized trials (which partly drew on the same registry) before the outcomes were extracted: GLP-1RAs and gallbladder or biliary disease (risk ratio (RR) 1.37), [31] SGLT2 inhibitors and ketoacidosis (odds ratio 2.13),[32] thiazolidinediones and heart failure (RR 1.72),[33] DOACs versus vitamin-K antagonists (VKAs) and intracranial haemorrhage (RR 0.48),[34] and SGLT2 inhibitors and hospitalization for heart failure (hazard ratio 0.69).[35] The data were judged able to detect modest effects on events that are not trial endpoints if at least two of the first three were recovered (95% CI excluding 1 in the expected direction); the last two, which are trial endpoints, were reported descriptively.

### Statistical analysis

Each comparison formed a 2×2 table of events and participants at risk; within a trial, comparisons of one class that shared a control arm, or an active arm, were merged. Class effects were pooled as Mantel–Haenszel RRs with Greenland–Robins variance.[36] The primary analysis used placebo- or usual-care-controlled trials with an adverse-event window of at least 24 weeks or no stated window. Sensitivity analyses used DerSimonian–Laird random effects, [37] Peto odds ratios, windows of at least 52 weeks, leave-one-out, maximum-term counting, division of shared controls, exclusion of low-confidence adjudications and of joint-disease populations, terms restricted to arthroplasty or to osteoarthritis, and (post hoc) the trials in the upper half of outcome capture.

Eight hypotheses concerned GLP-1RAs (H1), SGLT2 inhibitors (H2), DPP-4 inhibitors (H3), interleukin (IL)-1β inhibition or colchicine (H4), anti-NGF antibodies (H5), bone-active agents (H6), statins or proprotein convertase subtilisin/kexin type 9 (PCSK9) inhibitors (H7), and VKAs versus DOACs in head-to-head trials (H8). Holm-adjusted P values are reported as pre-specified. The protocol’s other head-to-head pairs (tirzepatide versus other GLP-1RAs; DPP-4 inhibitors versus sulfonylureas) were run in the second addendum. GLP-1RA effects were stratified by trial population and molecule and related to the adverse-event window by inverse-variance-weighted meta-regression (Supplementary Table S3); a within-trial comparison of the highest and lowest doses was added before any class-level result was computed (Supplementary Table S4). All classes with at least 10 osteoarthritis-related SAEs were screened with false discovery rate control (the drug-class atlas).

To allow for systematic error, we fitted an empirical null distribution to all class-level negative-control estimates with at least 10 events, shifted each log RR by the null mean and added the null variance. This extends P-value calibration[38] to intervals; it does not model error as a function of effect size, as full interval calibration does.[39] Because a few precise estimates dominate the null, we report calibrated results as a range across several nulls. The primary null used all estimates. The refitted null (first addendum) excluded pairs with a known or strong effect. The second addendum added three further nulls: one without corticosteroid–cataract pairs (a biological prior), one restricted to estimates with at least 50 events and, for each hypothesis, one without that class’s own estimates. For precision, we report the smallest RR detectable with 80% power, and we use 0.80 to 1.25 as a precision benchmark, not as a clinical margin. Sex-specific counts are not posted, so analyses by sex were not possible. Analyses used Python 3.11 (pandas, NumPy, SciPy, statsmodels). Reporting follows the PRISMA 2020 statement where applicable (Supplementary File 5).

### Comparison with observational studies

After the hypothesis analyses, we compared published observational estimates of hip or knee replacement for the pre-specified drug classes with the randomized estimates (post hoc). A first, targeted search (Europe PMC, to 27 September 2026) was replaced by a systematic search of PubMed and Europe PMC to 11 October 2026, with eligibility rules fixed before searching (second addendum). Eligible studies were observational and reported an adjusted ratio estimate with 95% CI for hip or knee replacement as a separate outcome, against non-use or an active comparator. A language model screened the records and extracted the estimates. Each extracted number was matched by script to a verbatim quote from its source (and, if taken from an abstract, to the PubMed record), and two authors (ZX and MP) jointly checked every extracted estimate against its source. We took the authors’ main adjusted estimate (when several models were reported, the one given in the abstract) per study, comparison, population and joint; secondary exposure definitions, indications and sensitivity analyses were listed but not counted. Active-comparator contrasts used adjusted indirect comparisons of the two placebo-controlled class estimates (Bucher method). We tested each difference on the log scale and classed each pair as differing (P < 0.05) or, if not, as compatible and informative (randomized 95% CI upper-to-lower limit ratio of at most 2) or uninformative (wider CI).

### Patient and public involvement

Patients and the public were not involved in the design, conduct, reporting or dissemination plans of this study.

## Results

### Trials and outcome capture

Of 5,240 trials, 3,940 were eligible, 1,699 had a protocol-outcome SAE and were adjudicated, and 1,429 contributed 1,673 randomized comparisons (Figure 1B; Supplementary Figure S1). The primary analysis comprised 881 placebo- or usual-care-controlled trials with 1,751,625 randomized participants and 9,408 osteoarthritis-related SAEs across 272 drug classes (Table 1). In the 868 trials that posted baseline sex, 41.7% of 1,790,749 participants were women (baseline counts include arms that were not compared). Median enrolment was 925. The median stated adverse-event window was 104 weeks (interquartile range 54 to 242), and 25% of trials did not state one (Figure 1D). Cardiovascular, renal and metabolic trials contributed a third of the trials and two thirds of the events (Figure 1C). “Osteoarthritis” accounted for 88% of event terms, explicit arthroplasty or replacement terms for 8%, and rapidly progressive osteoarthritis for 3%.

**Table 1.** Characteristics of the 881 trials in the primary analysis.

| Characteristic | Category | Trials, n (%) | Participants | OA-related SAEs |
| --- | --- | --- | --- | --- |
| Therapeutic area | Cardiovascular / renal / metabolic | 294 (33.4) | 1,009,144 | 6,377 |
| Therapeutic area | Oncology | 201 (22.8) | 214,778 | 497 |
| Therapeutic area | Other | 117 (13.3) | 189,468 | 805 |
| Therapeutic area | Musculoskeletal / pain | 113 (12.8) | 129,846 | 1,362 |
| Therapeutic area | Respiratory | 80 (9.1) | 130,117 | 140 |
| Therapeutic area | Neuro / psychiatric | 76 (8.6) | 78,272 | 227 |
| Phase | Phase 2 or 2/3 | 73 (8.3) | 65,312 | 225 |
| Phase | Phase 3 | 770 (87.4) | 1,585,925 | 8,612 |
| Phase | Phase 4 | 36 (4.1) | 98,242 | 570 |
| Phase | Other/not stated | 2 (0.2) | 2,146 | 1 |
| Sponsor | Industry | 836 (94.9) | 1,634,588 | 8,849 |
| Sponsor | Non-industry | 45 (5.1) | 117,037 | 559 |
| Completion year | 2002–2009 | 119 (13.5) | 200,281 | 1,334 |
| Completion year | 2010–2016 | 363 (41.2) | 722,106 | 3,763 |
| Completion year | 2017–2025 | 399 (45.3) | 829,238 | 4,311 |
| Adverse-event reporting window | 24–51 weeks | 100 (11.4) | 90,874 | 107 |
| Adverse-event reporting window | 52–103 weeks | 192 (21.8) | 255,466 | 897 |
| Adverse-event reporting window | ≥104 weeks | 367 (41.7) | 986,912 | 6,443 |
| Adverse-event reporting window | Not stated | 222 (25.2) | 418,373 | 1,961 |
| Control | Placebo | 791 (89.8) | 1,643,914 | 9,296 |
| Control | Usual care | 89 (10.1) | 106,280 | 110 |
| Control | Placebo and usual care | 1 (0.1) | 1,431 | 2 |
| Enrolment | 500–999 | 477 (54.1) | 313,309 | 656 |
| Enrolment | 1,000–4,999 | 320 (36.3) | 548,967 | 1,939 |
| Enrolment | ≥5,000 | 84 (9.5) | 889,349 | 6,813 |
| Population | Joint disease or joint pain | 71 (8.1) | 59,718 | 852 |
| Population | Other populations | 810 (91.9) | 1,691,907 | 8,556 |

Participants are those in the randomized arms that contributed to the primary comparisons. OA, osteoarthritis; SAE, serious adverse event. Therapeutic areas were assigned from registered conditions; “placebo and usual care” denotes a trial with both control types.

Capture of the outcome was incomplete and varied between trials. In the control arms of 70 large trials (at least 2,000 control participants and a stated window of at least two years; post hoc), the median rate was 1.4 osteoarthritis-related SAEs per 1,000 person-years (interquartile range 0.8 to 2.3). By comparison, 12% of Look AHEAD participants, adults with type 2 diabetes and overweight, reported a knee replacement over more than a decade.[18] In CANTOS, the registry placebo arm recorded 51 events, against about 67 replacements implied by the published rate (capture about 76%; Supplementary Table S12).

### The registry detects known drug effects

All three pre-specified positive controls were recovered (Figure 2A; Supplementary Figure S2), although the canakinumab and gout controls rest on trials in the preliminary analysis (98% and 94% of events). Canakinumab reduced osteoarthritis-related SAEs (59/7,410 vs 52/3,849; RR 0.57, 0.39 to 0.82), matching the hazard ratio of 0.58 in the CANTOS database.[15] Anti-NGF antibodies increased them (12 trials; RR 2.31, 1.82 to 2.92), and the excess persisted without the largest trial (1.55, 1.07 to 2.25) and with maximum-term counting (3.08, 2.19 to 4.34). The excess was coded as rapidly progressive osteoarthritis (RR 8.07, 4.41 to 14.77) and osteoarthritis (1.71, 1.15 to 2.53), not as arthroplasty (1.06, 0.73 to 1.52; three trials with events; Figure 2C; Supplementary Table S8). SGLT2 inhibitors reduced gout SAEs (46 trials, 16 with events; RR 0.66, 0.45 to 0.99), but the reduction was fragile (0.68, 0.45 to 1.01, with windows of at least 52 weeks; 0.66, 0.41 to 1.05, with maximum-term counting). The published estimate of 0.55 refers to a composite that included starting urate-lowering drugs.[30]

**Figure 2.**
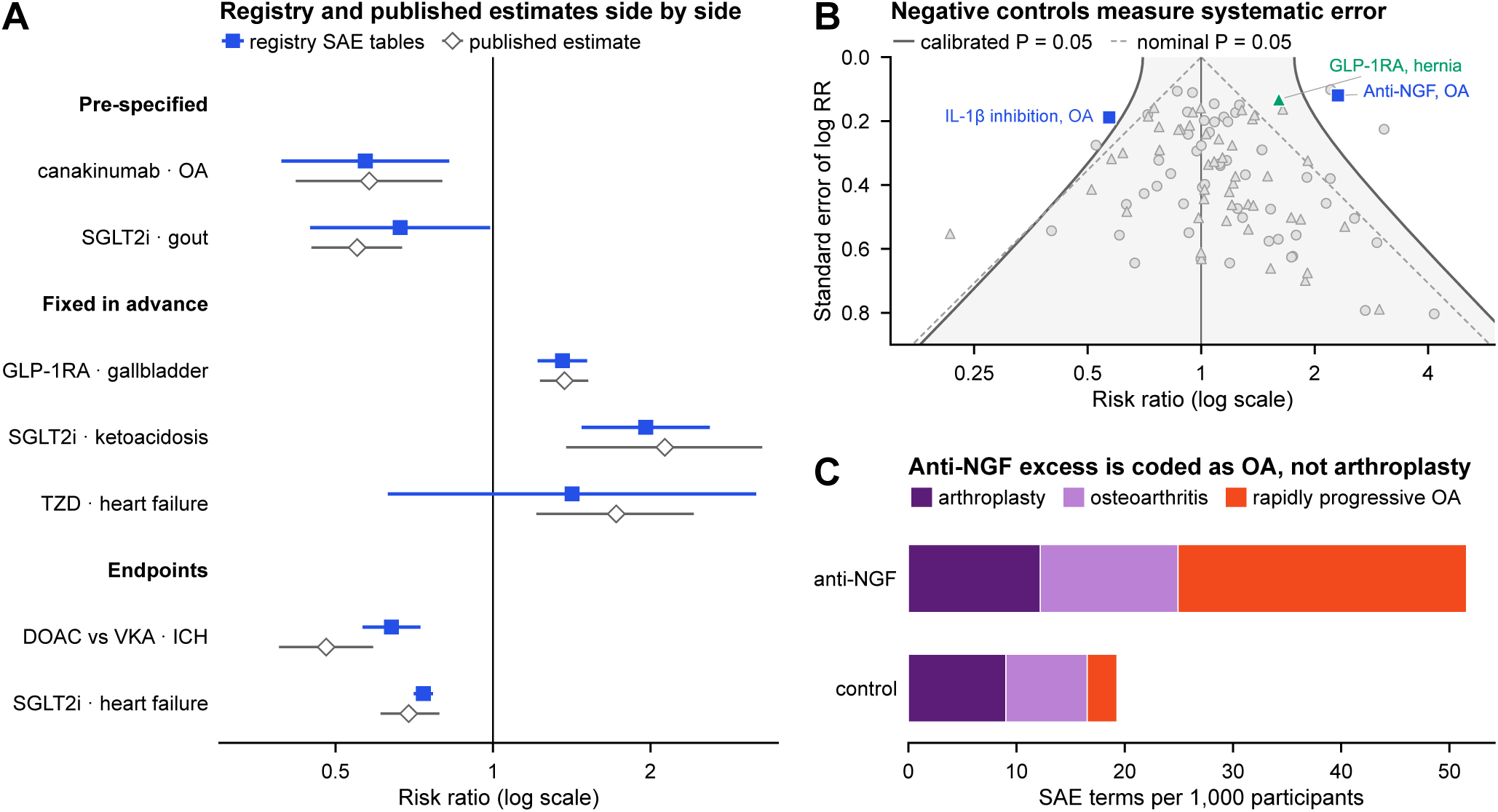
Validation with positive and negative controls. (A) Registry Mantel–Haenszel risk ratios (RR; filled squares, 95% CI) above published randomized estimates (open diamonds) for two pre-specified positive controls (canakinumab and osteoarthritis-related SAEs, from the same trial, CANTOS;[15] SGLT2 inhibitors and gout[30]), three controls whose expected size was fixed before extraction (published meta-analyses[31–33]) and two trial endpoints reported descriptively.[34, 35] Events and values: Supplementary Figure S2 and Supplementary Table S9. (B) Negative-control outcomes: class-level estimates for cataract (circles) and inguinal hernia (triangles) against their standard errors. Shaded area between the solid lines, not significant after empirical calibration (primary null: mean 0.11, SD 0.24 on the log scale, 100 estimates); dashed lines, nominal P = 0.05. Squares, osteoarthritis-related SAEs with anti-NGF antibodies and IL-1β inhibition; green triangle, GLP-1RAs and inguinal hernia (Supplementary Figure S6). (C) SAE terms per 1,000 participants in the 12 anti-NGF trials (crude pooled rates), by coded term; RR 8.07 (4.41 to 14.77) for rapidly progressive osteoarthritis, 1.71 (1.15 to 2.53) for osteoarthritis and 1.06 (0.73 to 1.52) for arthroplasty. ICH, intracranial haemorrhage; DOAC, direct oral anticoagulant; VKA, vitamin-K antagonist; GLP-1RA, glucagon-like peptide-1 receptor agonist; SGLT2i, sodium–glucose cotransporter-2 inhibitor; TZD, thiazolidinedione; NGF, nerve growth factor; IL, interleukin; OA, osteoarthritis; SAE, serious adverse event.

The modest-effect controls met the pre-stated criterion. Two of the three non-endpoint controls were recovered: GLP-1RAs and gallbladder or biliary disease (49 trials; RR 1.36, 1.22 to 1.51; expected 1.37) and SGLT2 inhibitors and ketoacidosis (1.96, 1.48 to 2.60; expected 2.13). The estimate for thiazolidinediones and heart failure (7 trials, 31 events; 1.42, 0.63 to 3.18) was too imprecise to judge (Figure 2A). Of the two trial-endpoint controls, heart failure with SGLT2 inhibitors (0.74, 0.71 to 0.77; expected 0.69) matched, while intracranial haemorrhage with DOACs versus VKAs (0.64, 0.56 to 0.73; expected 0.48) was attenuated (Supplementary Table S9).

The negative-control outcomes were not null, so the protocol’s expectation of null effects was not met. The primary empirical null had a mean of 0.11 and an SD of 0.24 on the log scale (100 estimates; Figure 2B), driven by a few precise estimates: corticosteroids and cataract, and inguinal hernia with GLP-1RAs (152 vs 89 events; RR 1.61, 1.24 to 2.08) and with PCSK9 inhibitors (1.65, 1.20 to 2.27; Supplementary Figure S6). Without the corticosteroid–cataract pairs, the null had a mean of 0.07 (SD 0.15). Class effects on osteoarthritis-related SAEs were not correlated with effects on cataract or hernia (inverse-variance-weighted r −0.01 and 0.01; Supplementary Figure S7).

### GLP-1 receptor agonists and SGLT2 inhibitors

In the same trials that showed their known effects, neither GLP-1RAs nor SGLT2 inhibitors detectably changed osteoarthritis-related SAEs (Figure 3A). In the 49 GLP-1RA trials (126,956 participants), the RR for osteoarthritis-related SAEs was 0.97 (0.86 to 1.10; 508 vs 491 events; I² = 0%); in the 46 SGLT2 inhibitor trials, it was 0.96 (0.82 to 1.12). In the trials with events in the upper half of outcome capture (post hoc), the estimates were 1.03 (0.90 to 1.19) for GLP-1RAs and 1.06 (0.86 to 1.31) for SGLT2 inhibitors. In these trials the effects on gallbladder disease (1.32), ketoacidosis (1.63) and heart failure (0.75) persisted, while the gout estimate lost precision (0.54, 0.26 to 1.14). After calibration, the GLP-1RA estimate was 0.87 (0.54 to 1.41) with the primary null and 0.92 (0.70 to 1.21) with the refitted null; the SGLT2 inhibitor estimates were 0.86 (0.53 to 1.40) and 0.91 (0.68 to 1.22), respectively (Supplementary Table S12). The smallest RR detectable with 80% power was 0.84 for GLP-1RAs and 0.80 for SGLT2 inhibitors nominally, and 0.46 to 0.67 across the calibrated nulls. Of the GLP-1RA events, 93% came from the 13 GLP-1RA trials in the preliminary analysis; the other 36 GLP-1RA trials gave 1.12 (0.67 to 1.88).

**Figure 3.**
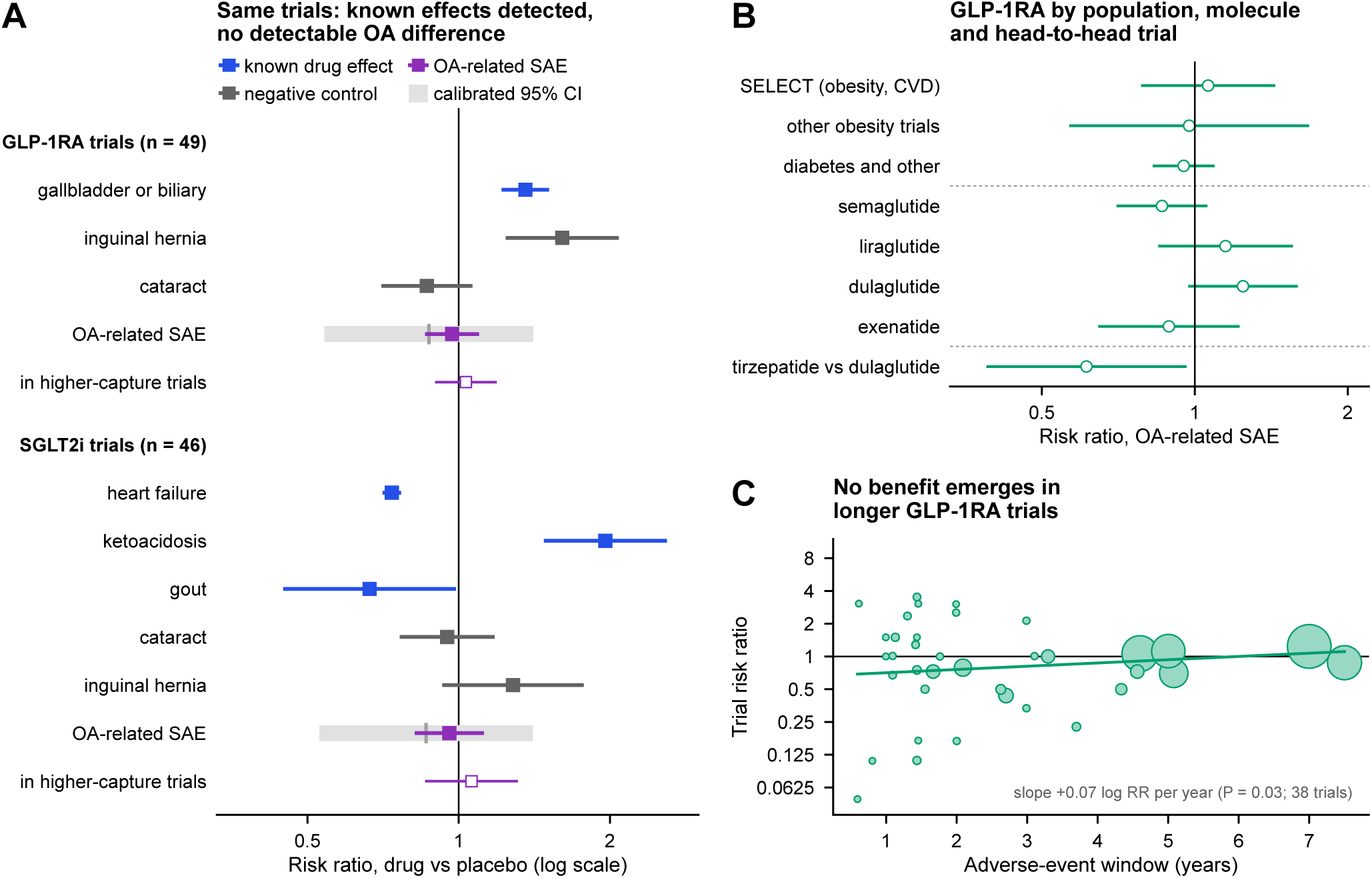
GLP-1 receptor agonists and SGLT2 inhibitors: known effects detected, no detectable difference in osteoarthritis-related SAEs. (A) Mantel–Haenszel risk ratios (95% CI) against placebo or usual care in the 49 GLP-1RA and 46 SGLT2 inhibitor trials of the primary analysis, for outcomes with known effects of these drugs (blue), negative-control outcomes (grey) and osteoarthritis-related SAEs (purple; grey bar and tick, calibrated 95% CI and estimate with the primary null; open square, trials in the upper half of outcome capture). GLP-1RAs: gallbladder 1.36 (1.22 to 1.51); osteoarthritis-related SAEs 0.97 (0.86 to 1.10), calibrated 0.87 (0.54 to 1.41). SGLT2 inhibitors: heart failure 0.74 (0.71 to 0.77), ketoacidosis 1.96 (1.48 to 2.60), gout 0.66 (0.45 to 0.99); osteoarthritis-related SAEs 0.96 (0.82 to 1.12), calibrated 0.86 (0.53 to 1.40). (B) GLP-1RAs by trial population (SELECT shown alone), by molecule (molecules with at least 100 events; heterogeneity across all nine molecules, P = 0.17) and in the head-to-head SURPASS-CVOT trial. (C) Trial risk ratios against the adverse-event window (circle area proportional to events; the 38 trials with at least one event and a stated window), with the inverse-variance-weighted meta-regression line. CVD, cardiovascular disease; GLP-1RA, glucagon-like peptide-1 receptor agonist; OA, osteoarthritis; SAE, serious adverse event; SGLT2i, sodium–glucose cotransporter-2 inhibitor.

No subgroup or molecule pointed consistently in one direction (Figure 3B). The estimate in obesity trials (22 trials; 1.04, 0.80 to 1.36) rested largely on SELECT (165 of 227 events; RR 1.06, 85/8,803 vs 80/8,801);[40] other obesity trials gave 0.97 (0.57 to 1.68), and diabetes and other trials 0.95 (0.83 to 1.09). Among molecules with at least 100 events, estimates ranged from 0.86 (0.70 to 1.06) for semaglutide to 1.24 (0.97 to 1.59) for dulaglutide (heterogeneity across all nine molecules, P = 0.17; Supplementary Table S11). In the one large head-to-head trial, SURPASS-CVOT, tirzepatide had fewer osteoarthritis-related SAEs than dulaglutide (30/6,647 vs 49/6,647; RR 0.61, 0.39 to 0.96). Longer trials did not show more benefit: trial estimates rose slightly with the adverse-event window (+0.07 log RR per year, P = 0.03, 38 trials; Figure 3C; Supplementary Figure S5 and Table S3).

### Other proposed drugs and the full atlas

For the other proposed drugs, estimates were less precise and, apart from IL-1β inhibition and anti-NGF antibodies, compatible with no effect (Figure 4A). DPP-4 inhibitors gave 1.26 (0.99 to 1.61; Holm-adjusted P = 0.38), almost entirely from the trials of the preliminary analysis (1.28, 0.99 to 1.65); the other trials were uninformative (1.12, 0.49 to 2.53), and linagliptin versus glimepiride in CAROLINA gave 0.81 (0.57 to 1.16). The bone-active estimate (13 trials; 1.00, 0.81 to 1.24) came mostly from denosumab trials (304 of 336 events; 1.03, 0.83 to 1.29); bisphosphonate trials contributed only 11 events. PCSK9 inhibitors (1.09, 0.93 to 1.27) supplied 626 of the 665 events in H7; four statin trials gave 0.95 (0.51 to 1.78). In 21 head-to-head trials, VKAs versus DOACs gave 1.10 (0.92 to 1.30), 1.09 (0.92 to 1.31) in atrial fibrillation trials and 1.12 (0.94 to 1.34) in trials whose arms received the same antiplatelet therapy (post hoc). IL-1β inhibition (0.57, 0.39 to 0.82; Holm-adjusted P = 0.020) rested on CANTOS, and no colchicine trial contributed events. The conclusions for H1–H8 did not change in the sensitivity analyses or after calibration (Supplementary Tables S1, S10 and S12).

**Figure 4.**
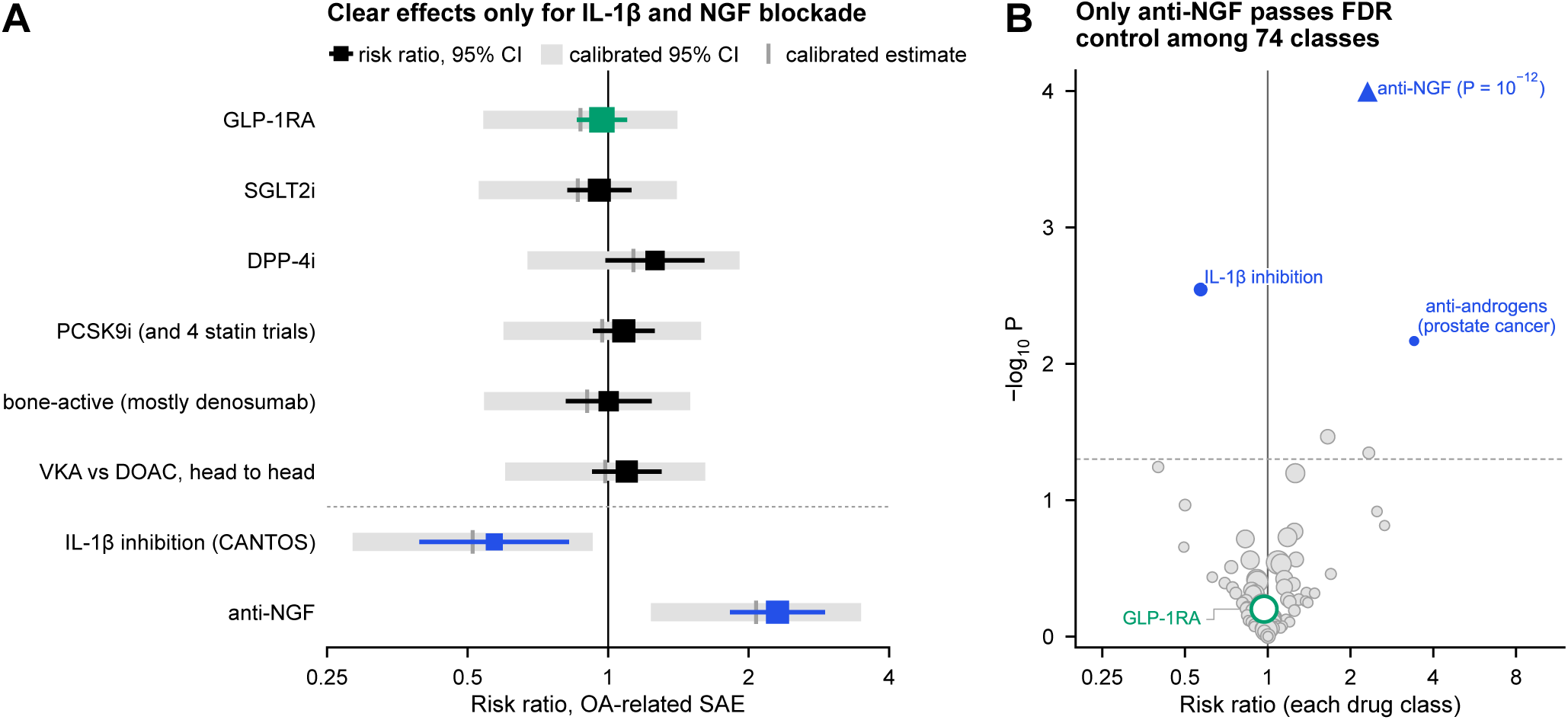
Other pre-specified hypotheses and the drug-class atlas. (A) Mantel–Haenszel risk ratios with 95% CIs for hypotheses H1–H8 (marker area increases with events; green, GLP-1RAs; blue, classes significant after calibration; IL-1β inhibition rests on CANTOS); grey bars and ticks, calibrated 95% CIs and estimates (primary null). DPP-4i 1.26 (0.99 to 1.61); statins and PCSK9 inhibitors (H7) 1.08 (0.93 to 1.26); bone-active agents, mostly denosumab, 1.00 (0.81 to 1.24); VKA versus DOAC, head to head, 1.10 (0.92 to 1.30); IL-1β inhibition 0.57 (0.39 to 0.82); anti-NGF 2.31 (1.82 to 2.92). Trials, events, Holm-adjusted P values and estimates outside the preliminary analysis: Supplementary Figure S3. (B) Risk ratio against −log₁₀ P for the 74 drug classes with at least 10 osteoarthritis-related SAEs (marker area increases with events; blue, significant after calibration with the primary null; dashed line, P = 0.05). Anti-NGF antibodies (triangle) lie above the axis limit (−log₁₀ P = 11.6) and are the only class passing false discovery rate control (q < 0.001); all classes are listed in Supplementary Table S2. DOAC, direct oral anticoagulant; DPP-4i, dipeptidyl peptidase-4 inhibitor; FDR, false discovery rate; GLP-1RA, glucagon-like peptide-1 receptor agonist; IL, interleukin; NGF, nerve growth factor; OA, osteoarthritis; PCSK9i, proprotein convertase subtilisin/kexin type 9 inhibitor; SAE, serious adverse event; SGLT2i, sodium–glucose cotransporter-2 inhibitor; VKA, vitamin-K antagonist.

Across the drug-class atlas of 74 classes with at least 10 osteoarthritis-related SAEs, only anti-NGF antibodies passed false discovery rate control (q < 0.001; Figure 4B; Supplementary Table S2). Anti-androgens (8 trials; 25 vs 6 events; 3.41, 1.40 to 8.29), vitamin D (2 trials; 1.65, 1.04 to 2.63) and inhaled anticholinergics (17 trials; 27 events; 2.33, 1.02 to 5.34) were nominally significant. After calibration, anti-NGF antibodies (calibrated P = 0.006), IL-1β inhibition (0.027) and anti-androgens (0.028) remained significant. All anti-androgen trials were in prostate cancer, in which these drugs prolonged survival (deaths RR 0.82, 0.77 to 0.88) and time on treatment, and so the period of adverse-event collection. Only 4 of the 74 classes had 95% CIs within 0.80 to 1.25.

### Observational versus randomized estimates

The systematic search found 18 eligible studies (7,282 records retrieved; 3,487 unique records screened), and 21 estimates from 16 studies were compared (Figure 5; Supplementary Figure S4 and Table S6). Eight estimates, from 5 studies, differed from the randomized estimates at P < 0.05, in both directions: for GLP-1RAs, hazard ratios of 0.60 and 0.75 in people with obesity and hip or knee osteoarthritis and 2.61 (knee) in those without osteoarthritis (randomized, 1.04 in obesity trials);[3] for SGLT2 inhibitors, 0.70 versus GLP-1RAs[5] and 1.33 versus non-use;[41] for DPP-4 inhibitors, 0.38 and 0.42 (randomized, 1.26);[7] and for warfarin versus DOACs, an odds ratio of 1.59 (randomized, 1.10).[8] Six estimates were compatible and informative, among them the two most recent GLP-1RA cohorts (0.90 and 0.91)[42, 43] and three of the four SGLT2 inhibitor versus GLP-1RA estimates.[6, 44] Seven were uninformative because the randomized data for bisphosphonates, denosumab and statins were sparse.[10–12] The count depended on the analysis choices (Figure 5B): 9 estimates differed with alternative comparators, and 3 (primary null) or 6 (refitted null) after the randomized intervals were widened for systematic error. Two estimates whose direction could not be read from the source were not counted; when oriented as implied by the absolute risks reported with them, both were compatible.[6]

**Figure 5.**
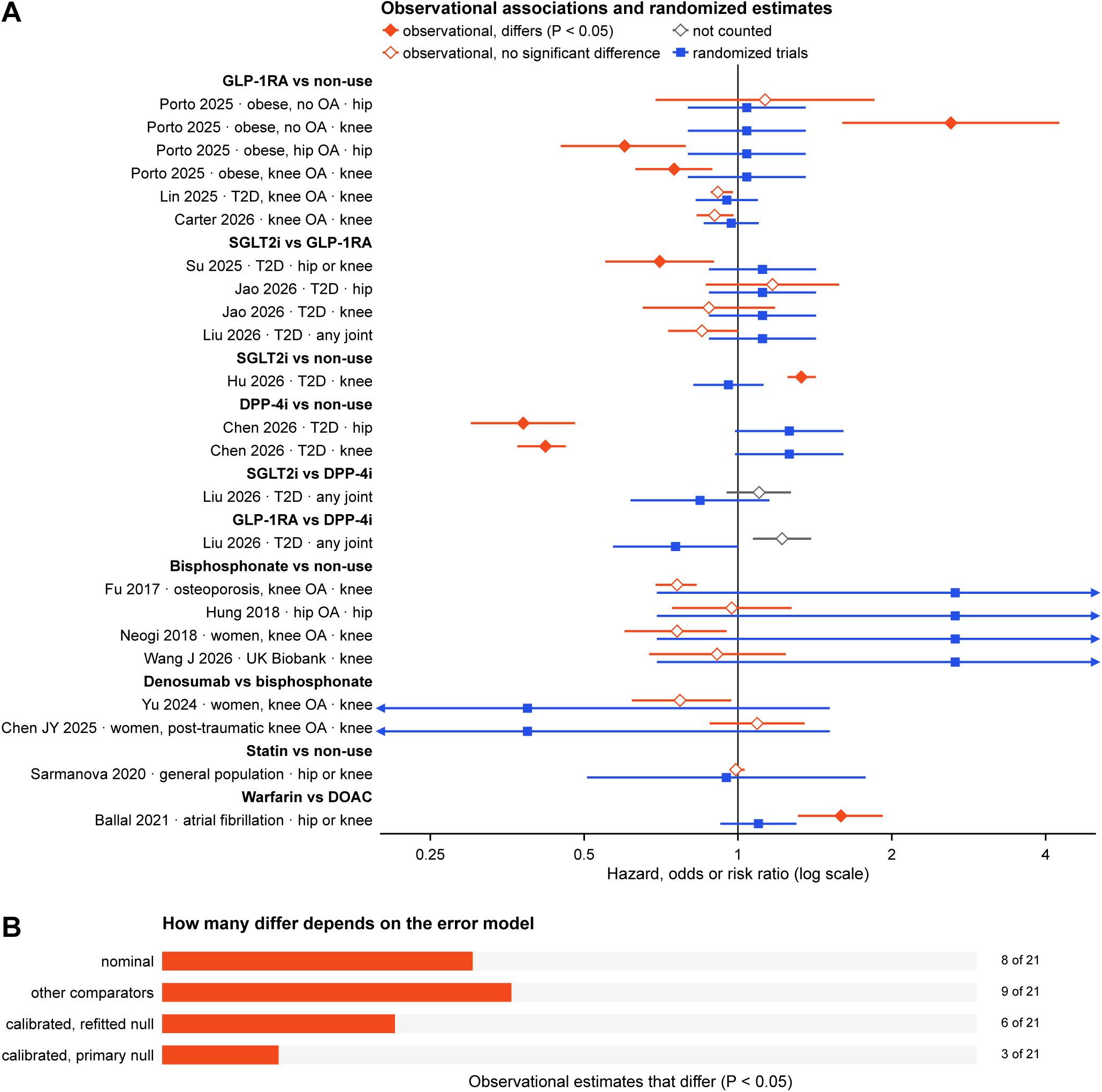
Published observational associations against randomized estimates for the same drug classes. (A) For each eligible study from a systematic search of PubMed and Europe PMC to 11 October 2026 (eligibility fixed before searching), the authors’ main estimate per comparison, population and joint: diamonds, adjusted hazard or odds ratio (95% CI) for hip or knee replacement; squares, the randomized Mantel–Haenszel risk ratio (95% CI) for the same contrast (GLP-1RA trials by population; bisphosphonate trials for bisphosphonates; adjusted indirect comparisons for active comparators). Filled diamonds, difference at P < 0.05 (test of the difference between log estimates); grey diamonds, two estimates whose direction cannot be read from the source (not counted); arrows, intervals beyond the axis. (B) Number of the 21 counted estimates that differ with the primary comparators, with alternative comparators (bone-active agents for bisphosphonates, DPP-4 inhibitor trials outside the preliminary analysis, all SGLT2 inhibitor trials), and after widening the randomized intervals for systematic error with the refitted or the primary empirical null. Values, sources and the 14 further eligible estimates listed but not counted: Supplementary Figure S4 and Supplementary Table S6. DOAC, direct oral anticoagulant; DPP-4i, dipeptidyl peptidase-4 inhibitor; GLP-1RA, glucagon-like peptide-1 receptor agonist; OA, osteoarthritis; SGLT2i, sodium–glucose cotransporter-2 inhibitor; T2D, type 2 diabetes.

## Discussion

Posted SAE tables of 881 randomized trials detected known drug effects, including modest ones. In the same GLP-1RA and SGLT2 inhibitor trials that showed those drugs’ effects on gallbladder disease, ketoacidosis, heart failure and gout, osteoarthritis-related SAEs did not differ detectably. Nominal 95% CIs excluded reductions larger than 14–18%, but calibrated intervals could not exclude reductions of up to 30–32% (refitted null) or 46–47% (primary null). For most other proposed drugs the data were compatible with no effect but too imprecise to exclude relevant effects. Of 21 published observational estimates for the same drug classes, 8 differed from the randomized estimates, in both directions, and 3 to 6 did after allowing for systematic error.

### What the registry can and cannot detect

The central question for any analysis of registry SAE tables is whether they record true drug effects on the outcome of interest. The canakinumab control cannot answer it, because CANTOS ascertained replacements from the same database,[15] although it suggests that the registry recorded about three quarters of replacements in that trial. The modest-effect controls show that acute, drug-caused admissions are recorded well enough to detect modest effects, in the same trials and reporting systems that produced the osteoarthritis estimates. They do not show the same for elective surgery. Capture of osteoarthritis admissions was incomplete and varied, at about 1 to 2 per 1,000 person-years in large outcome trials, whereas 12% of Look AHEAD participants reported a knee replacement over more than a decade.[18] Incomplete capture that is equal in both arms costs precision but does not bias the RR; bias arises if admissions unrelated to replacement are counted, or if capture differs between arms. The null estimates held in the trials with the highest capture.

The anti-NGF findings show what a term-level breakdown can and cannot say. The arthroplasty-coded estimate (1.06) agrees with placebo-controlled data with adjudicated joint outcomes, in which joint replacement was not significantly increased (5.9–7.0% vs 4.5%; RR 1.13, 0.76 to 1.68),[45, 46] whereas the excess was clearer against non-steroidal anti-inflammatory drugs (NSAIDs) and with fasinumab.[19, 20, 47] In the NSAID-controlled tanezumab trial, no SAE in either arm was coded as arthroplasty, which suggests that replacements may be recorded under the condition that led to them. A breakdown by term therefore cannot separate disease progression from surgery.

### GLP-1 receptor agonists and SGLT2 inhibitors

The hope that GLP-1RAs will modify osteoarthritis rests on weight loss, symptomatic benefit in STEP 9,[27] cohort data[48] and a gut–joint mechanism in mice.[49] In randomized comparisons of about 127,000 participants, GLP-1RAs did not detectably reduce osteoarthritis-related SAEs. This agrees with meta-analyses of osteoarthritis adverse events[21, 22] and with a target-trial emulation of incident osteoarthritis.[50]

Three biases could distort these estimates, and the data speak to each. First, many trials stop collecting adverse events soon after the last dose, and GLP-1RA users discontinue more often; this shortens observation in active arms and would bias towards apparent benefit, not towards the null. Second, weight loss can make patients eligible for surgery, which would bias towards harm. The excess of inguinal-hernia admissions with GLP-1RAs, plausibly from vomiting, constipation and weight loss, suggests that admissions for elective surgery do respond to these drugs. Third, benefit may take longer than the trials ran, but estimates did not fall as windows lengthened. Randomized weight loss by other means points the same way: lifestyle intervention in Look AHEAD did not reduce knee replacement overall,[18] and centrally acting anti-obesity drugs gave 0.99 (0.83 to 1.18) in our data.

The evidence in obesity comes essentially from SELECT, in people with cardiovascular disease, so a benefit confined to established knee osteoarthritis, or to semaglutide (0.86, 0.70 to 1.06), cannot be excluded. Tirzepatide had fewer events than dulaglutide in one trial from the preliminary analysis; as the only large randomized comparison of two incretin drugs that differ in weight loss, this finding deserves follow-up.

### Other drugs

For DPP-4 inhibitors, the estimate leaned towards harm but came almost entirely from the trials of the preliminary analysis that motivated the hypothesis; the other trials were too sparse to confirm or refute it. The bone-active estimate is essentially a denosumab estimate; a zoledronic acid trial found more knee replacements during treatment,[17] and the bisphosphonate data here are too sparse to add to it. Statin data are uninformative, and PCSK9 inhibitors showed no clear effect. The warfarin hypothesis[8, 9] is not refuted: head-to-head trials are compatible with an excess of up to 30% (34% in trials whose arms received the same antiplatelet therapy; upper limits 41–74% after calibration), surgery is harder to schedule in patients taking a VKA, and most trials ran for about two years. Colchicine, perhaps the most promising repurposing candidate, could not be assessed, because LoDoCo2[16] is registered outside ClinicalTrials.gov.

### Why observational estimates disagree

Observational estimates disagreed with each other as much as with the trials. For GLP-1RAs they were protective in people with obesity and osteoarthritis, harmful in those without it, and close to null in the two most recent cohorts; for SGLT2 inhibitors they were protective relative to GLP-1RAs in one cohort and harmful relative to non-use in another.[3, 5, 41–43] Confounding by indication and channelling, healthy-adherer effects and differential access to surgery could each produce such discrepancies. Our comparison was post hoc and compared different estimands: SAEs in trials, mostly in people without diagnosed osteoarthritis, against arthroplasty in routine care, often in people with it. The number of discrepant estimates also depended on the comparator and on the model of systematic error (Figure 5B). A population-specific effect, for example one confined to established osteoarthritis, could be real and still differ from these trials. The comparison is best read as showing that the large protective associations reported for DPP-4 inhibitors, and for GLP-1RAs in osteoarthritis, were not reproduced in randomized data, rather than as a test of each study.

### Lessons for registry analyses

Negative-control outcomes need not be null. Inguinal hernia, chosen as unrelated to drug biology, was more frequent with GLP-1RAs and PCSK9 inhibitors, and corticosteroids increased cataract. A few such pairs determined both the shift and the width of the calibration, so calibrated results are best read as a range, and negative controls should be screened for known effects before they are used. Unexpected signals need similar caution: the anti-androgen excess persisted when trials reported over unequal periods were removed, but it may still reflect longer on-treatment observation of active arms in prostate cancer, or musculoskeletal effects of more complete androgen blockade.

### Strengths and limitations

Strengths include the scale of the randomized data, positive controls of large and modest size tested in the same trials as the hypotheses, negative-control outcomes, calibration, estimates reported separately for trials outside the preliminary analysis, and full disclosure of deviations and corrected errors. The study also has limitations. The analysis plan was written after a preliminary pooled analysis that covered most events of several hypotheses, and it was not registered. The modest-effect controls were chosen after the main results were known, and the code implementing their term lists was committed together with their results. Arm adjudication by a language model followed written rules but was masked by instruction only. An automated re-audit, followed by screens of all trials, found rules misapplied in 102 of the 1,499 trials analysed before correction (7%); these were corrected. The human check of a 202-trial sample was done jointly by two authors, not independently in duplicate. The outcome is an incompletely captured proxy for joint replacement that was not clinically adjudicated, and summed term counts overstate precision where participants have several terms. Observation was short, adverse-event collection often ended soon after treatment, and posting is incomplete for older trials. IL-1β inhibition rests on one trial, colchicine was absent, and most classes were informative only about large effects.

## Conclusions

Posted SAE tables of randomized trials detected known drug effects, including modest ones. In trials that detected their known effects, GLP-1 receptor agonists and SGLT2 inhibitors had no detectable effect on osteoarthritis-related SAEs over about two years, and several large associations reported in observational studies were not reproduced. Registry analyses can screen repurposing candidates for large, short-term effects before trials are funded; slow structural effects need dedicated trials.

## Declarations

## Contributors

ZX, XG and XZ conceived the study. ZX designed the study and its analysis rules, performed the analyses, prepared the figures and wrote the manuscript. ZX and MP checked the trial classification, the extracted estimates, the key results and the references. JC, XC and JL contributed to the interpretation of the results and critically revised the manuscript. XG and XZ supervised the study, acquired funding and critically revised the manuscript. All authors reviewed and approved the final manuscript. ZX is the guarantor, accepts full responsibility for the work and the conduct of the study, had access to the data and controlled the decision to publish. *Use of artificial intelligence:* the authors posed the research question, set the direction of the study, approved its design, selected the methods and results to report, checked the trial classification, the extracted estimates, the key results and the references, and revised and approved the manuscript. Under their direction, a large language model (Claude Opus 5.5, Anthropic) classified the trial arms, screened the literature search and extracted the published estimates (see Methods), wrote and ran the analysis code, reviewed drafts and assisted with drafting and editing. The authors take full responsibility for the content.

## Funding

This work was supported by the Scientific Research Project of the Health Commission of Hubei Province (WJ2023F086) and the 2024 Key Project of the Department of Science and Technology of Hubei Province (2024BCB059). The funders had no role in study design, data collection, analysis and interpretation, writing of the report or the decision to submit the paper for publication.

## Competing interests

None declared.

## Ethics approval

Not required. The study used only publicly available, aggregate trial results; no individual-participant data were accessed.

## Patient consent for publication

Not applicable.

## Data availability statement

All data relevant to the study are included in the article or uploaded as supplementary information. The source records are public on ClinicalTrials.gov.[26] Trial identifiers, adjudication records, extracted counts and all class-level results are in the Supplementary Data. The analysis code, adjudication rules and search records are provided to the editors and reviewers with this submission and will be deposited in a public repository with a DOI upon publication.

## Related work

The same registry source was used by the authors in a separately submitted analysis of common, non-serious adverse events in placebo-controlled trials (Xu Z, Cheng J, Chen X, et al., submitted), which addresses a different question and outcome.

## Notes

### Competing Interest Statement

The authors have declared no competing interest.

### Author Declarations

The study used ONLY openly available, aggregate data: (1) trial results posted on ClinicalTrials.gov (https://clinicaltrials.gov/), downloaded through the ClinicalTrials.gov API version 2 (https://clinicaltrials.gov/data-api/api) on 5 October 2026 and restricted to trials whose results were first posted by 26 September 2026; and (2) effect estimates from published observational studies, identified through PubMed (https://pubmed.ncbi.nlm.nih.gov/) and Europe PMC (https://europepmc.org/) and cited in the manuscript. No individual-participant data were used.

