## Supplementary Appendix for "Do drugs proposed to protect or harm joints change serious osteoarthritis events? A meta-analysis of adverse event tables from 881 randomized trials"

#### Contents

- Supplementary Methods
  - S1. Protocol, timeline and the preliminary pooled analysis
  - S2. Data source and eligibility
  - S3. Arm adjudication
  - S4. Outcome definitions
  - S5. Drug classes
  - S6. Statistical methods
  - S7. Deviations from the protocol and corrections
  - S8. Comparison with published observational estimates
  - S9. Reporting checklist
- Supplementary Tables S1–S12
- Supplementary Figures S1–S7
- Separate files:
  - Supplementary Data (Excel workbook with all class-level results, trial lists and the trials of the preliminary analysis);
  - Supplementary File 1 (the protocol, verbatim);
  - Supplementary File 2 (the adjudication rules, verbatim);
  - Supplementary File 3 (the horizon-scan note that contains the preliminary pooled analysis, verbatim; partly in Chinese), with an English translation of the Chinese passages;
  - Supplementary File 4 (Protocol addendum D7, verbatim);
  - Supplementary File 5 (PRISMA 2020 checklist);
  - Supplementary File 6 (Protocol addendum D9, verbatim);
  - Supplementary File 7 (Protocol addendum D10, verbatim).

### Supplementary Methods

#### S1. Protocol, timeline and the preliminary pooled analysis

**Protocol.** The protocol (Supplementary File 1) specified eligibility, outcome term lists, pre-specified mechanistic classes, eight hypotheses with directions, the positive-control criterion (at least two of three recovered), the negative-control outcomes, the primary estimator, sensitivity analyses and multiplicity handling. It was time-stamped with a SHA-256 hash in a version-controlled repository and was not registered externally.

**The preliminary pooled analysis.** While candidate research questions were being screened, a horizon-scan note (Supplementary File 3) extracted SAE tables of 72 large trials from the same registry and pooled them with Mantel–Haenszel risk ratios. The protocol describes this as a look at "about 60" trials; the list it refers to (Supplementary Data, sheet S9) contains 72. The note reported, among others:

| Contrast in the preliminary analysis | Events, active vs control | RR (95% CI) | Corresponding analysis |
|---|---|---|---|
| Canakinumab (CANTOS) | 58/6,718 vs 51/3,348 | 0.57 (0.39–0.82) | P1, H4 |
| DPP-4 inhibitors (5 outcome trials) | 133/23,833 vs 104/23,750 | 1.28 (0.99–1.65) | H3 |
| GLP-1RAs (11 trials, including SELECT) | 445/46,055 vs 455/44,727 | 0.97 (0.85–1.11) | H1 |
| SGLT2 inhibitors (13 trials) | 321/47,841 vs 280/41,275 | 0.93 (0.79–1.09) | H2 |
| Warfarin vs DOACs (4 atrial-fibrillation trials) | 200/29,223 vs 258/42,219 | 1.12 (0.93–1.35) | H8 |
| Denosumab (FREEDOM); romosozumab (FRAME) | 63/3,886 vs 79/3,876; 6/3,581 vs 15/3,576 | 0.80; 0.40 | H6 |
| PCSK9 antibodies (FOURIER, ODYSSEY) | 144/23,220 vs 146/23,199 | 0.99 | H7 |

The note also identified the observational studies of warfarin and DPP-4 inhibitors that were later set against the randomized estimates. Supplementary Table S7 gives, for every positive control and hypothesis, the share of informative events from these 72 trials and the estimate from the other trials. For P1/H4, P3, H1, H2, H3 and H8 the share is 87–98%, so these analyses re-estimate results that were known when the protocol was written; only the anti-NGF analyses (7% of events previewed) and, in part, H6 and H7 (about half) are largely independent of the preliminary analysis. The manuscript therefore treats the hypothesis tests as estimation.

**Timeline (UTC, 26 September 2026; repository history).** Horizon-scan note with the preliminary analysis committed 10:38; protocol committed 11:46; adjudication of 1,699 trials and the drug-class map frozen 12:41; first class-level results committed 12:45. The window-parser correction (D4) followed at 16:31 after internal review. Corrections C1–C9 and the analyses of the addendum (D7) were made on 10 October 2026 (US Pacific time; 11 October UTC) after a second review; the addendum was committed (5da2c6d, refined in 077328a) before those analyses were run. Corrections C10–C11 and analyses R1–R7 (addendum D9, Supplementary File 6) followed on 11 October 2026 after two further simulated peer reviews; the addendum was committed (f5255bb) before they were run. On 12 October 2026 the verification workbook came back with its human-verifier columns empty and with verdicts added by an automated audit (a language model); its flags were checked against the registry records, and corrections C12–C18, screens of all included trials for the same errors and analysis R8 followed (addendum D10, Supplementary File 7, committed 042fde3 before they were applied; screen S4 was defined afterwards, S3). All review rounds were carried out by language models acting as internal reviewers, methodologists, clinicians and editors (simulated review); no external human review has taken place.

#### S2. Data source and eligibility

**Source.** The original query of the ClinicalTrials.gov API (version 2) on 26 September 2026 returned 5,240 interventional, randomized trials with an enrolment of at least 500, posted results and at least one drug or biological intervention. The corpus used here was rebuilt from a complete download of all studies with posted results made on 5 October 2026, restricted by the same criteria and to trials whose results were first posted by 26 September 2026; it contains the same 5,240 trials. Records edited between the two dates carry their edits; the only material edit (NCT04736199) does not affect any reported estimate.

**Automatic exclusions** (1,300 trials; categories overlap): minimum age below 16 years; vaccine trials; crossover, sequential or single-group designs; populations defined by orthopaedic, arthroplasty or fracture surgery.

**Trials not adjudicated.** Trials with no osteoarthritis-related, gout, cataract or inguinal-hernia SAE in any reporting group (2,241 trials) contribute no information to Mantel–Haenszel estimates and were not adjudicated. The modest-effect positive controls (S6) were therefore estimated within the 1,699 adjudicated trials; this sample is incomplete for gallbladder disease, ketoacidosis, heart failure and intracranial haemorrhage, which also occur in trials without any of the four protocol outcomes.

#### S3. Arm adjudication

**Inputs.** For each of the 1,699 informative trials, a structured card listed title, conditions, phase, minimum age, the adverse-event time-frame text, protocol arms and interventions, and the adverse-event reporting groups (identifier, title, description, number at risk) with a rule-based suggested role and drug name. Cards contained no adverse-event counts.

**Rules** (Supplementary File 2). Only randomized core-period groups are retained; totals, open-label extensions, run-in periods and non-randomized cohorts are excluded. Doses of the same drug are pooled. Add-on designs are placebo comparisons of the added drug. Several active drugs sharing one control each form a comparison with that control. Active-comparator trials are head-to-head comparisons. Factorial trials are analysed by margins when the groups allow. Strategy trials without a specific added drug are excluded. Each decision carries a confidence rating.

**Adjudicator.** Adjudication was carried out in eight parallel batches by a large language model (Claude Opus 5.5; Anthropic), working as an agent in the analysis environment, given the rules and the cards and instructed not to use outcome data. The outcome tables were present in the shared working directory, so masking to outcomes was by instruction, not by access control; the model may also carry background knowledge of published trial results, and it named some trials (for example FINESSE and ZFAST) in its comments. The protocol (section 4) had specified a rule-based classification with manual review of ambiguous groups; adjudication of every trial by the model is a deviation (S7, D8).

**Structural validation.** Every card was adjudicated exactly once; all group identifiers existed; no group was both active and control; every included trial had at least one comparison. Corrections after review are listed in S7 (C5, C6).

**Confidence.** Of the 1,429 trials analysed after corrections C7 and C12–C17 (20 further trials with randomized comparisons lacked posted denominators), adjudication confidence was high for 1,252, medium for 170 and low for 7.

**Re-audit and screens (addendum D10).** An automated audit of the 202-trial verification sample flagged 24 trials; the registry records confirmed misapplied rules in 14 (C12–C17), and the screens below confirmed one more (MSB11456 vs tocilizumab). The same error types were then sought in every included trial with four text screens (S1, nested records and extensions; S2, groups mixing randomized arms or switched inside the adverse-event window; S3, active or strategy comparators and VKA comparators; S4, groups reported over different periods, added after S1–S3 had missed such cases). The 1,120 hits in 732 trials were read against the registry text by language-model reviewers without outcome counts, every kept hit involving a switch, crossover, escape, extension or re-randomization was re-checked under one rule, and the analyst (the language model that ran the analysis) decided the few unresolved hits. A comparison was dropped when a compared group included participants randomized to another arm, when one arm received the other arm's treatment inside the window without separate reporting, or when the active group was reported beyond the end of the control group's reporting period (for example, placebo to week 16 against the drug to week 52, or a placebo arm cut at a crossover after unblinding while the active arm continued in open-label extension); censoring of individual non-responders at an early-escape visit within a common placebo-controlled period, as-treated reassignment of a few misdosed participants and differences in planned treatment length between regimens were kept. Two further errors of the same kinds were found incidentally during the review. In all, 14 trials were excluded, 86 comparisons in 69 trials were dropped, and the comparisons of 23 trials were re-specified or restored; 102 of the 1,499 previously analysed trials were affected; every decision, with the registry text it rests on, is in the repository (`adjudication/audit_D10.tsv`). The human check below covered the corrected classification of the 202 sampled trials, not every screen decision.

**Human verification.** Two authors (ZX and MP), working together, checked a stratified sample of 202 trials (verification workbook) against the registry records, without looking at adverse-event counts: (A) every trial with at least 5 events in a positive-control or hypothesis analysis (77 trials); (B) the 20 primary-analysis trials with the most osteoarthritis-related SAEs; (C) a random sample of 100 trials with randomized comparisons (seed 2026); (D) 20 random trials excluded by the adjudicator. The check was made on 12 October 2026, after the corrections of addendum D10, which the workbook showed next to each sampled trial. They confirmed the inclusion decision, the group assignment and the drug under test for all 202 trials (100% for each; Wilson 95% CI 98.1 to 100; by stratum 77/77, 20/20, 100/100 and 20/20). The planned independent duplicate check was not done (the two authors worked jointly), so agreement between verifiers (κ) cannot be reported. The verdicts of the automated audit (D10) are not human verification and are not counted here.

### S4. Outcome definitions

Outcomes were defined on posted serious-event terms, matched case-insensitively.

**Osteoarthritis-related SAE** (called "joint-failure SAE" in the protocol). A term was counted if it matched `osteoarthritis | osteoarthrosis | arthrosis | arthroplasty | joint replacement | knee replacement | hip replacement | shoulder replacement | joint prosthesis implantation | rapidly progressive osteoarthritis | chondrolysis` and did not match `spin | vertebr | cervical | lumbar | thoracic | intervertebral | facet | jaw | infect | septic |`

`bacterial | loosening | dislocation | failure of | revision | complication | fracture | periprosthetic | haemarth | hemarth | pseudarth | pseudoarth | traumatic arthrosis` (the last five stems were added before analysis, D1). Hand, foot and unspecified osteoarthritis terms are included; "large joint" in the protocol was not enforced by the term list.

**Counting.** Within a reporting group, affected participants were summed across matching terms; the maximum across terms (a participant count) is a sensitivity analysis.

**Term composition** (primary set, 9,408 events): osteoarthritis 88.0%; knee arthroplasty 3.7%; rapidly progressive osteoarthritis 3.3%; hip arthroplasty 2.8%; other replacement or arthroplasty terms 1.7%; other osteoarthritis terms 0.5%. Arthroplasty or replacement terms together: 8.1%.

**Capture of the outcome** (addendum D9, R2). SAE definitions follow ICH E2A, and some protocols exempt admissions planned before enrolment, so whether an elective replacement is reported as an SAE depends on the sponsor, and a replacement is usually coded under the condition that led to it (in the NSAID-controlled tanezumab trial NCT02528188, 40 vs 4 osteoarthritis-related SAEs were posted and none was coded as arthroplasty). Capture was estimated as control-arm events per 1,000 person-years, taking the stated adverse-event window as follow-up: in 70 placebo- or usual-care-controlled trials with at least 2,000 control participants and a stated window of at least 104 weeks, the median was 1.38 (interquartile range 0.80 to 2.29; pooled 1.79); by therapeutic area, pooled rates ranged from 0.44 (oncology) to 2.34 (musculoskeletal and pain). In CANTOS, 51 placebo-arm events stood against about 67 replacements implied by the published placebo rate (0.54 per 100 person-years over a median 3.7 years in 3,344 participants), a capture of 76% (Supplementary Table S12b). Pre-specified estimates were repeated in trials at or above the median combined-arm rate (as defined in the addendum) and, because that definition mainly removed trials without events, in the upper half of trials with at least one event (added after the first; Supplementary Table S12c–d).

**Other outcomes.** Gout SAE: `gout | gouty`. Cataract SAE (negative control): `cataract`. Inguinal-hernia SAE (negative control): `inguinal hernia`. Spinal degenerative SAE and osteonecrosis SAE (secondary; Supplementary Data). Deaths: per-group death counts.

**Outcomes added in addendum D7.** Arthroplasty or replacement terms only; osteoarthritis terms only (rapidly progressive osteoarthritis and chondrolysis excluded); rapidly progressive osteoarthritis or chondrolysis; an anti-NGF joint-safety composite (osteoarthritis-related terms, osteonecrosis other than of the jaw, subchondral insufficiency fracture); and four outcomes for the modest-effect positive controls, as implemented in `02_build_tables.py`: gallbladder or biliary disease (`cholelithiasis | cholecystitis | gall ?bladder | biliary colic | choledocholithiasis | cholangitis | bile duct stone | biliary obstruction`, excluding `cancer | carcinoma | neoplasm | malignan | polyp`); ketoacidosis (`ketoacidosis | ketosis`); heart failure (`cardiac failure | heart failure | ventricular failure | cardiac decompensation`); and intracranial haemorrhage (`intracranial | cerebral | cerebellar | subdural | subarachnoid | intraventricular | extradural | epidural | brain stem | thalamic | basal ganglia | putamen` followed by `haemorrhage | hemorrhage | haematoma | hematoma | bleed`, or `haemorrhagic stroke | haemorrhage intracranial | haemorrhagic cerebral infarction | haemorrhagic transformation`, in either spelling).

### S5. Drug classes

Active drugs were recorded by international non-proprietary name and mapped to pre-specified mechanistic classes (GLP-1RAs including dual and triple agonists; SGLT2 inhibitors; DPP-4 inhibitors; IL-1 inhibitors; colchicine; anti-NGF antibodies; bisphosphonates, denosumab, romosozumab; statins; PCSK9 inhibitors; vitamin-K antagonists; DOACs; thiazolidinediones; other protocol classes), otherwise to the fourth ATC level through NLM RxNav (first ATC code returned), or labelled as unclassified single agents. For three drugs the first code returned was that of a combination partner (dutasteride, tadalafil, netarsudil); they were reassigned to their own codes (C10). Fixed-dose combinations were classified by the component that differed between arms. Corrections C3 (cagrilintide, an amylin analogue, is its own class) and C4 (vitamin D3 and cholecalciferol form one class) were applied after review. ATC-derived classes can mix routes: ATC D07AC contains inhaled and intranasal fluticasone and mometasone; ATC A01AC contains intravitreal triamcinolone and systemic dexamethasone. The protocol's manual mechanistic mapping of investigational drugs was not done (D8); odanacatib and sprifermin remain unclassified single agents.

### S6. Statistical methods

**Comparison tables and pooling.** Each comparison gave a 2×2 table (a, c: participants with the event in the active and control arms; $n_1$, $n_0$: participants at risk). Comparisons of one class sharing a control arm within a trial were merged, and so were comparisons in which the same active group was compared with several control groups of the same class (C10). The Mantel–Haenszel RR, $\Sigma(a_i n_{0i}/N_i)/\Sigma(c_i n_{1i}/N_i)$, has Greenland–Robins variance; trials with no event carry no weight. DerSimonian–Laird random effects (0.5 continuity correction) and Peto odds ratios are sensitivity analyses.

**Multiplicity.** Two-sided P values for H1–H8 were Holm-adjusted; the atlas screen used Benjamini–Hochberg FDR across classes with at least 10 osteoarthritis-related SAEs.

**Empirical calibration.** A normal systematic-error distribution was fitted by maximum likelihood to the log RRs of all class-level negative-control estimates with at least 10 events (100 estimates; mean 0.106, SD 0.236), accounting for each estimate's standard error. Calibrated estimates shift each log RR by the null mean and add the null variance to its variance; calibrated P values and CIs follow from these. This extends the P-value calibration of Schuemie and colleagues to intervals; it does not implement their interval calibration, which models systematic error as a function of the true effect using synthetic positive controls. Because the dispersion of the null comes mainly from a few high-information pairs, calibration is reported as a bracket over five nulls: the four of addendum D9 R4 (all estimates; without corticosteroid–cataract pairs, defined biologically, not by P value, 97 estimates, mean 0.066, SD 0.152; estimates with at least 50 events, 33, 0.087, 0.267; and, for each hypothesis, a leave-one-class-out null) and the refitted null of addendum D7 A3 (without known-effect estimates; 96, 0.051, 0.126) (Supplementary Table S12f–g). Addendum A3 refitted the null without the estimates with a known pharmacological cause or nominal $P < 0.001$ (four after D10: corticosteroids of three routes and cataract; GLP-1RAs and inguinal hernia) and to each negative-control outcome alone (Supplementary Table S10).

**Precision.** The smallest RR detectable with 80% power at two-sided $\alpha = 0.05$ is $\exp(\pm 2.80 \times SE)$ for the observed SE of the log RR (Supplementary Tables S2 and S11).

**Secondary and sensitivity analyses** (pre-specified unless stated). Competing mortality: denominators rescaled to $N \times (1 - ½ \times \text{proportion dying})$ per arm; deaths were posted for 519 of 988 primary comparisons, and the others were left unadjusted. Duration: inverse-variance-weighted meta-regression of trial log RR on the adverse-event window (the protocol named median follow-up, which the registry does not record; D8). GLP-1RA population strata: obesity trials were those whose conditions or title mention obesity, overweight, weight management or weight loss. Dose (D2b, corrected by C8): within-trial highest vs lowest randomized dose of the drug under test. Added before results (D2a, D3, D6): division of shared control arms; exclusion of low-confidence adjudications; exclusion of joint-disease populations. Addendum D7: estimates outside the preliminary analysis (A1), outcome construct (A2), calibration (A3), precision (A4), GLP-1RA by molecule with Cochran's Q (A5), modest-effect positive controls (A6).

**Analysis of addendum D10 (post hoc, defined before it was run).** R8: H8 and the intracranial-haemorrhage control without the trials that assign different antiplatelet co-therapy by arm (PIONEER AF-PCI, RE-DUAL PCI, ENTRUST-AF PCI, PETRO; Supplementary Table S5e).

**Analyses of addendum D9 (post hoc, defined before they were run).** R1: the protocol's head-to-head pairs (tirzepatide vs other GLP-1RAs; DPP-4 inhibitors vs sulfonylureas). R2: capture of the outcome (S4). R3: composition of the hypothesis estimates (SELECT and other obesity trials; denosumab, bisphosphonates and romosozumab; PCSK9 inhibitors and statins; anti-NGF without the largest trial and with maximum-term counting; VKA vs DOAC in atrial-fibrillation trials). R4: calibration bracket (above). R5: estimates for the H1 and H2 trial sets on all outcomes (Figure 3A). R6–R7: systematic search for observational estimates and comparison rules (S8).

**Modest-effect positive controls (A6).** Expected effects and the success criterion were committed before extraction (Supplementary File 4). Analysis sets were those of the hypotheses: placebo or usual-care trials with windows of at least 24 weeks or not stated, and DOAC vs VKA head-to-head trials (shown as DOAC vs VKA).

**Software.** Python 3.11 (pandas 2.3, NumPy 2.4, SciPy 1.17, statsmodels 0.15). A shuffled-label dummy mode tested the code before the real analysis. Figures were drawn with matplotlib.

#### S7. Deviations from the protocol and corrections

Deviations D1–D3 were made before any class-level result was computed.

**D1. Outcome definition.** The stem "arthrosis" matched haemarthrosis, pseudarthrosis and traumatic arthrosis; these were excluded after a corpus-wide term-frequency audit made without reference to arm or drug.

**D2. Two sensitivity analyses added** for consistency with a companion analysis: (a) equal division of control counts among classes sharing a control arm; (b) the within-trial dose comparison. While the dose code was tested on the rule-based pre-classification with shuffled labels, unshuffled high-vs-low dose estimates were displayed for three classes (anti-NGF 0.66, SGLT2 inhibitors 0.72, romosozumab 0.21); no primary or hypothesis estimate was displayed, and the dummy mode was corrected.

**D3. Exclusion of low-confidence adjudications** (sensitivity analysis), added after adjudicators reported delayed-start and rescue designs with shorter control windows.

**D4. Correction of the adverse-event window parser** after the first results (26 September 2026). The original parser missed unit-first ("Week 52"), interposed-word, hyphenated and range formats and kept short matches such as "+ 1 day", so some long trials were classed as shorter than 24 weeks. The corrected parser changed the window status of 181 trials (into the primary window) and 284 (out of it). Pre-specified estimates changed little (Supplementary Table S5a); an apparent excess in ATC D07AC (inhaled or intranasal corticosteroids; RR 8.24 → 1.06) disappeared.

**D5. Protocol items not performed or not reported.** "Any musculoskeletal SAE" was not extracted; head-to-head comparisons other than VKA vs DOAC were not run (the protocol's two other pairs were run in addendum D9, R1); person-years are not reported for the estimates because 25% of trials do not state an adverse-event window (the capture analysis of addendum D9 approximates them from stated windows); secondary outcomes (spinal degenerative disease, osteonecrosis) are in Supplementary Data without interpretation.

**D6. Departures found in a first audit of the manuscript** (none affects a conclusion): the age criterion admitted two trials with minimum age 16–17 years and a stated maximum; the joint-disease-population sensitivity analysis had not been logged; the calibration null used estimates with at least 10 events; the ≥24-week window had not been applied to H8 (now corrected, C2) and the ≥52-week restriction had not been applied to P2 (recomputed in Supplementary Table S1).

**D7. Corrections and additional analyses after a second internal review** (Protocol addendum, Supplementary File 4; before/after estimates in Supplementary Table S5b):

- C1: offsets such as "through 30 days after the last dose" had still been read as the adverse-event window (for example SCOUT, STABILITY, VOYAGER-PAD); offsets of at most 3 months are now removed before parsing, and 196 trials changed from shorter than 24 weeks to 24 weeks or not stated. The primary set grew from 908 to 945 trials and from 8,872 to 9,610 events.
- C2: eight DOAC-vs-VKA trials whose comparator was written "vitamin K antagonist" or a combination were missing from H8; H8 now has 20 trials within the protocol window (23 in all windows).
- C3: cagrilintide reclassified as an amylin analogue. C4: vitamin D3 and cholecalciferol merged.

- C5: three glycaemic sub-studies of VERTIS-CV, whose participants belong to the cardiovascular-outcome population, removed. C6: the placebo comparison of AWARD-5, whose placebo group switched to sitagliptin, removed.
- C7: comparisons without posted denominators removed (20 trials). C8: the dose analysis de-duplicated, restricted to the primary window and reading the dose of the drug under test. C9: missing deaths no longer treated as zero.
- A1–A7: post hoc analyses defined in the addendum before they were run (S6, S8). The term lists implemented for the gallbladder and intracranial-haemorrhage controls were written at the same time as the addendum and before extraction, but are slightly broader than its wording (biliary obstruction; extradural, epidural, brain-stem, thalamic, basal-ganglia and putamen haemorrhage; haemorrhagic cerebral infarction or transformation; neoplasms and polyps excluded) (S4).

No pre-specified conclusion changed (Supplementary Table S5b).

**D8. Departures not logged at the time** (recorded after the second review): adjudication of every trial by a language model instead of rule-based classification with manual review of ambiguous groups (protocol section 4); no manual mechanistic mapping of investigational drugs (section 5); meta-regression on the adverse-event window instead of median follow-up (section 9); class-level negative-control flags, now reported in Supplementary Table S2, and death RRs, in Supplementary Data; the exploratory vaccine class was not analysed because vaccine trials were excluded; the size of the preliminary analysis ("about 60" vs 72 trials, S1).

**D9. Corrections and additional analyses after two simulated peer reviews** (Protocol addendum D9, Supplementary File 6, committed f5255bb before the analyses were run; before/after estimates in Supplementary Table S5c):

- C10: in four combination-versus-monotherapy trials the combination arm entered one class estimate twice; three drugs had been mapped to a combination partner's ATC class (S5). Active groups now enter a class estimate once.
- C11: on-treatment adverse-event windows with an offset (for example "up to and including 7 days following the date of last dose") had still been read as windows of 0–1 week in multi-year trials such as EUCLID. All 87 analysable trials with a parsed window under 24 weeks and either ≥1,000 participants or ≥1 protocol-outcome event were read individually by the language model that performed the analysis (not by a human; two authors later checked all 87 decisions, as revised under D10, and agreed with them); 19 windows defined as treatment duration plus an offset were set to not stated (list with reasons in the repository).

The primary set grew from 945 to 953 trials and from 9,610 to 9,615 events; H8 gained two trials (1.12, 0.95 to 1.33 → 1.12, 0.94 to 1.32); no other pre-specified estimate changed.

- R1–R7: post hoc analyses (S6, S8). One variant, the capture sensitivity analysis restricted to trials with at least one event, was added after the addendum's own definition proved uninformative and is labelled as such.

**D10. Corrections after an automated re-audit of the verification workbook** (Protocol addendum D10, Supplementary File 7, committed 042fde3 before the corrections were applied; before/after estimates in Supplementary Table S5d):

- C12: EBBINGHAUS, a sub-study of FOURIER whose SAE table re-reports FOURIER data, excluded (the PCSK9 participants had been counted twice).
- C13: comparisons whose groups mixed randomized arms or were reported over different periods dropped (for example, tanezumab in chronic low back pain, whose tanezumab groups included placebo patients switched after week 16).
- C14: extension records (TULIP-LTE, PT003008 and others found by the screens) excluded.
- C15: comparison type corrected (lenalidomide added to rituximab; active or physician's-choice comparators now head-to-head; strategy trials such as the SECURE polypill trial excluded).
- C16: GALILEO's comparator is an antiplatelet strategy, not a VKA; the trial left H8 and the intracranial-haemorrhage control. KSI-301 renamed tarcocimab tedromer.
- C17: two parallel trials wrongly excluded as crossover were included (one adds no data, lacking posted denominators).
- C18: nine adverse-event windows corrected after the full time-frame text was read (the hand review of D9 had seen text cut at 300 characters), and the window of the restored study 603A (C17) set to its 12 weeks; NCT00441350, listed in the addendum, was kept at 8 weeks because only its first phase is compared.
- Screens S1–S4 of all included trials (S3); S4 was added after S1–S3, and its rule (reporting periods) was applied to every kept switch-related hit. One reviewer's report of VERIFY (vildagliptin) was decided by the analyst with the published design, because the registry does not describe the replacement of placebo by vildagliptin after treatment failure.
- R8: post hoc analysis (S6). The observational comparison now takes, when a study reports several models, the estimate given in its abstract (Chen JY 2025: inverse-probability-weighted 1.09, 0.88 to 1.35, instead of 1.04, 0.73 to 1.47); the class of the pair did not change.

The primary set fell from 953 to 881 trials and from 9,615 to 9,408 events (74 trials left: 51 whose placebo or usual-care comparisons were dropped, 12 excluded, 11 now head-to-head; 2 entered). H1, H2, H6 and H7 were unchanged to two decimals; anti-NGF 2.34 → 2.31; DPP-4 inhibitors 1.22 → 1.26 (VERIFY removed); VKA vs DOAC 1.12 → 1.10. No pre-specified conclusion changed; the DPP-4 inhibitor estimate outside the preliminary analysis moved from 0.90 (0.44 to 1.83) to 1.12 (0.49 to 2.53). In the atlas, the anti-androgen estimate rose (2.02 → 3.41) after the removal of prostate cancer trials whose placebo arms were cut at a crossover, and became significant after calibration with the primary null; inhaled anticholinergics became nominally significant (2.33, 1.02 to 5.34). The protocol's validation aim of null negative-control outcomes was not met (S6; Results).

### S8. Comparison with published observational estimates (post hoc)

**First, targeted search (addendum D7, superseded).** Europe PMC was searched on 27 September 2026 with queries combining each pre-specified class with "joint replacement" or "arthroplasty" and "osteoarthritis", ranked by citation count; the warfarin and DPP-4 inhibitor studies had been noted in the preliminary analysis (S1). It gave 11 counted estimates from 7 studies, of which 8 differed from the randomized estimates. Simulated peer review found that it missed eligible studies, so addendum D9 replaced it with a systematic search; the earlier comparison is kept in Supplementary Data (sheet S6c).

**Systematic search (addendum D9, R6; eligibility fixed before searching).** PubMed (MEDLINE) and Europe PMC were searched from inception to 11 October 2026, one query per drug class (GLP-1 receptor agonists including multi-agonists; SGLT2 inhibitors; DPP-4 inhibitors; IL-1 inhibitors or colchicine; anti-NGF antibodies; bisphosphonates; denosumab; romosozumab; statins; PCSK9 inhibitors; vitamin-K antagonists, for comparisons with DOACs) combined with replacement or arthroplasty terms; English, peer-reviewed, no preprints. Eligible were observational studies in humans reporting an adjusted or matched ratio estimate with 95% CI for hip or knee replacement as a separate outcome, against non-use or an active comparator; composite outcomes, Mendelian randomization, trials and reviews were excluded, and the reference lists of 35 relevant reviews were sought (31 available) and screened. Query strings, hit counts, every screening decision and the flow are in the repository (`03_analysis/B1_rct_atlas/obs_search_D9/`: searches.tsv, screening.tsv, flow.md).

**Flow.** 7,282 hits (PubMed 3,411; Europe PMC 3,871); 3,487 unique records screened by title and abstract (3,466 excluded; reasons assigned by a fixed rule hierarchy, so the counts by reason are approximate: no replacement outcome 1,235, review or meta-analysis 852, trial 570, not observational 436, other publication types 266, wrong exposure 96, no ratio estimate 10, Mendelian randomization 1); 16 further records from reference lists and 3 known candidates assessed; 26 full texts or, where no full text was open, abstracts assessed; 8 excluded (no replacement outcome 3, composite outcome 3, no ratio estimate 2); 18 studies eligible. No eligible observational study was found for IL-1 inhibitors, anti-NGF antibodies, romosozumab or PCSK9 inhibitors.

**Screening and extraction.** Screening and extraction were done by a language model, which recorded for every estimate a verbatim quote (at most 30 words) from the source; a script confirmed that each number appears in its quote, and every number taken from an abstract was matched again to the PubMed record. Two authors (ZX and MP) jointly checked all 37 extracted estimates against their sources (value, author's main model and direction) and confirmed them. For each study the authors' main estimate per comparison, population and joint was used (when several models were reported, the one given in the abstract; addendum D10); secondary exposure definitions (for example semaglutide or tirzepatide for 3 years), outcomes by indication, joint-specific sensitivity analyses and population subgroups were listed but not counted (Supplementary Table S6b). One eligible study (Lavu 2024) reports its hazard ratios only in a closed full text; raloxifene versus bisphosphonates was excluded because raloxifene is not a pre-specified class, and colchicine versus non-use has no randomized comparator.

**Comparison (R7).** The observational SE was (ln upper − ln lower)/3.92. Randomized comparators: GLP-1RA trials by population (obesity trials for obese populations; diabetes and other trials for type 2 diabetes; all trials otherwise); bisphosphonate trials for bisphosphonates (bone-active agents as sensitivity); denosumab versus bisphosphonates and SGLT2 inhibitors versus GLP-1RAs or DPP-4 inhibitors by adjusted indirect comparison (Bucher), with SGLT2 inhibitor trials outside heart-failure and kidney populations (all SGLT2 inhibitor trials as sensitivity); DPP-4 inhibitor trials (trials outside the preliminary analysis as sensitivity); VKA versus DOAC from H8. Differences were tested with $z = (\ln \text{obs} - \ln \text{rand})/\sqrt{(SE^2\text{obs} + SE^2\text{rand})}$, and the ratio of the two estimates is reported with its 95% CI. Each pair was classed as differing ($P < 0.05$), compatible and informative ($P \geq 0.05$ and randomized upper/lower CI ratio ≤ 2) or uninformative. Randomized intervals were also widened for systematic error with the empirical null (variance plus $SD^2$, shifted by the null mean for direct contrasts; plus 2 $SD^2$ without shift for indirect contrasts). Liu 2026 reports hazard ratios above 1 with lower absolute risks in the first-named group for both contrasts with DPP-4 inhibitors; these are not counted, and are oriented as the absolute risks imply in a sensitivity analysis.

**Result.** Twenty-one estimates from 16 studies were counted: 8, from 5 studies, differed at P < 0.05 (9 with the sensitivity comparators; 3 and 6 after calibration with the primary and refitted nulls); 6 were compatible and informative and 7 uninformative. With the two Liu 2026 contrasts oriented, 8 of 23 differed. The comparison is descriptive.

#### S9. Reporting checklist

The PRISMA 2020 checklist (Supplementary File 5) maps each item to the manuscript and this appendix. Items that concern study-level risk of bias, certainty of evidence and protocol registration are answered as not done, with reasons.

### Supplementary Table S1. Pre-specified sensitivity analyses (risk ratio, 95% CI)

### (a)

| Analysis | Primary (MH) | Random effects (DL) | Peto OR | Window ≥52 wk | Max-term count |
|---|---|---|---|---|---|
| P1 Canakinumab → OA-related SAE | 0.57 (0.39–0.82) | 0.57 (0.39–0.82) | 0.54 (0.36–0.81) | 0.57 (0.39–0.82) | 0.57 (0.39–0.82) |
| P2 Anti-NGF → OA-related SAE | 2.31 (1.82–2.92) | 1.87 (1.32–2.66) | 2.02 (1.67–2.44) | 2.43 (1.90–3.12) | 3.08 (2.19–4.34) |
| P3 SGLT2i → gout | 0.66 (0.45–0.99) | 0.66 (0.44–0.99) | 0.67 (0.45–0.98) | 0.68 (0.45–1.01) | 0.66 (0.41–1.05) |
| H1 GLP-1RA | 0.97 (0.86–1.10) | 0.96 (0.85–1.09) | 0.97 (0.85–1.10) | 0.97 (0.86–1.10) | 0.98 (0.86–1.11) |
| H1b GLP-1RA, outside the preliminary analysis | 1.12 (0.67–1.88) | 0.91 (0.55–1.48) | 1.12 (0.67–1.88) | 1.23 (0.71–2.14) | 1.18 (0.70–2.00) |
| H2 SGLT2i | 0.96 (0.82–1.12) | 0.95 (0.81–1.11) | 0.96 (0.82–1.12) | 0.98 (0.84–1.15) | 0.99 (0.84–1.16) |
| H3 DPP-4i | 1.26 (0.99–1.61) | 1.21 (0.95–1.55) | 1.26 (0.99–1.61) | 1.26 (0.99–1.61) | 1.26 (0.99–1.61) |
| H3b DPP-4i, outside the preliminary analysis | 1.12 (0.49–2.53) | 0.83 (0.40–1.72) | 1.12 (0.50–2.48) | 1.12 (0.49–2.53) | 1.12 (0.49–2.53) |
| H4 IL-1β inhibition | 0.57 (0.39–0.82) | 0.57 (0.39–0.82) | 0.54 (0.36–0.81) | 0.57 (0.39–0.82) | 0.57 (0.39–0.82) |
| H5 Anti-NGF | 2.31 (1.82–2.92) | 1.87 (1.32–2.66) | 2.02 (1.67–2.44) | 2.43 (1.90–3.12) | 3.08 (2.19–4.34) |
| H6 Bone-active agents | 1.00 (0.81–1.24) | 1.14 (0.78–1.67) | 1.00 (0.81–1.24) | 1.00 (0.81–1.24) | 0.98 (0.79–1.21) |
| H7 Statins + PCSK9i | 1.08 (0.93–1.26) | 1.08 (0.93–1.26) | 1.08 (0.93–1.26) | 1.11 (0.95–1.30) | 1.08 (0.92–1.25) |
| H8 VKA vs DOAC (head-to-head) | 1.10 (0.92–1.30) | 1.11 (0.93–1.31) | 1.10 (0.92–1.30) | 1.10 (0.93–1.31) | 1.10 (0.92–1.31) |
| H8b VKA vs DOAC, all windows | 1.10 (0.92–1.30) | 1.11 (0.93–1.32) | 1.10 (0.92–1.30) | 1.10 (0.92–1.30) | 1.10 (0.92–1.31) |

### (b)

| Analysis | Shared controls split | Excl. low-confidence | Excl. joint-disease populations | Leave-one-out range |
|---|---|---|---|---|
| P1 Canakinumab → OA-related SAE | 0.57 (0.39–0.82) | 0.57 (0.39–0.82) | 0.57 (0.39–0.82) | 0.56–0.74 |
| P2 Anti-NGF → OA-related SAE | 2.33 (1.84–2.94) | 2.31 (1.82–2.92) | not estimable* | 1.55–2.47 |
| P3 SGLT2i → gout | 0.66 (0.45–0.99) | 0.66 (0.45–0.99) | 0.66 (0.45–0.99) | 0.60–0.72 |
| H1 GLP-1RA | 0.97 (0.86–1.10) | 0.97 (0.86–1.10) | 0.97 (0.86–1.10) | 0.89–1.01 |
| H1b GLP-1RA, outside the preliminary analysis | 1.14 (0.67–1.93) | 1.12 (0.67–1.88) | 1.12 (0.67–1.88) | 1.05–1.32 |

| | | | | |
|---|---|---|---|---|
| H2 SGLT2i | 0.96 (0.82–1.12) | 0.96 (0.82–1.12) | 0.96 (0.82–1.12) | 0.89–0.98 |
| H3 DPP-4i | 1.27 (0.99–1.62) | 1.26 (0.99–1.61) | 1.26 (0.99–1.61) | 1.16–1.33 |
| H3b DPP-4i, outside the preliminary analysis | 1.22 (0.53–2.82) | 1.12 (0.49–2.53) | 1.12 (0.49–2.53) | 1.00–1.29 |
| H4 IL-1β inhibition | 0.57 (0.39–0.82) | 0.57 (0.39–0.82) | 0.57 (0.39–0.82) | 0.56–0.74 |
| H5 Anti-NGF | 2.33 (1.84–2.94) | 2.31 (1.82–2.92) | not estimable* | 1.55–2.47 |
| H6 Bone-active agents | 1.00 (0.81–1.24) | 1.00 (0.81–1.24) | 1.00 (0.81–1.23) | 0.96–1.19 |
| H7 Statins + PCSK9i | 1.08 (0.93–1.26) | 1.08 (0.93–1.26) | 1.08 (0.93–1.26) | 1.00–1.15 |
| H8 VKA vs DOAC (head-to-head) | 1.10 (0.92–1.30) | 1.10 (0.92–1.30) | 1.10 (0.92–1.30) | 1.03–1.18 |
| H8b VKA vs DOAC, all windows | 1.10 (0.92–1.30) | 1.10 (0.92–1.30) | 1.10 (0.92–1.30) | 1.03–1.17 |

*All anti-NGF trials with control-arm events enrolled patients with osteoarthritis or chronic pain. Leave-one-out ranges show point estimates only; omitting CANTOS from P1/H4 leaves two small trials (RR 0.74 (0.07–7.47)), so the IL-1β result rests on CANTOS. H8 applies the protocol window (≥24 weeks or not stated; correction C2); H8b includes head-to-head trials of any window. The ≥52-week value for P2 was recomputed because the analysis script had not applied the window restriction to it (deviation D6). H4a (colchicine) is omitted: the two eligible placebo-controlled colchicine trials recorded no protocol-outcome SAE in either arm. The sensitivity analyses for shared controls, low-confidence adjudications and joint-disease populations were added before results were computed (deviations D2, D3, D6). MH, Mantel–Haenszel; DL, DerSimonian–Laird.

### Supplementary Table S2. All drug classes with ≥10 OA-related SAEs (primary analysis)

| Class | Trials | Events (A vs C) | RR (95% CI) | P | FDR q | Calibrated RR (95% CI) | Calibrated P | Negative controls: cataract RR; hernia RR | Detectable RR (80% power) |
|---|---|---|---|---|---|---|---|---|---|
| Anti-NGF | 12 | 584 vs 77 | 2.31 (1.82–2.92) | 2.8e-12 | 2.1e-10 | 2.07 (1.23–3.48) | 0.006 | 0.32; 0.64 | 0.72 / 1.40 |
| IL-1 inhibitor | 3 | 59 vs 52 | 0.57 (0.39–0.82) | 0.003 | 0.11 | 0.51 (0.28–0.93) | 0.027 | 0.83; 1.22 | 0.59 / 1.69 |
| Anti-androgens (ATC L02BB) | 8 | 25 vs 6 | 3.41 (1.40–8.29) | 0.007 | 0.17 | 3.06 (1.13–8.34) | 0.028 | 4.15; 1.17 | 0.28 / 3.56 |
| Vitamin D and analogues (ATC A11CC) | 2 | 46 vs 27 | 1.65 (1.04–2.63) | 0.034 | 0.63 | 1.49 (0.77–2.86) | 0.24 | 0.00; — | 0.51 / 1.94 |
| Anticholinergics (ATC R03BB) | 17 | 19 vs 8 | 2.33 (1.02–5.34) | 0.045 | 0.67 | 2.10 (0.81–5.41) | 0.13 | 0.93; 1.33 | 0.31 / 3.27 |
| Romosozumab | 1 | 6 vs 15 | 0.40 (0.16–1.03) | 0.057 | 0.67 | 0.36 (0.13–1.03) | 0.057 | 1.00; — | 0.26 / 3.86 |
| DPP-4i | 30 | 150 vs 114 | 1.26 (0.99–1.61) | 0.063 | 0.67 | 1.13 (0.67–1.91) | 0.64 | 0.92; 1.11 | 0.71 / 1.42 |
| Atrasentan | 1 | 8 vs 16 | 0.50 (0.21–1.17) | 0.11 | 0.96 | 0.45 (0.17–1.18) | 0.10 | 2.20*; 1.50 | 0.30 / 3.35 |

| | | | | | | | | | |
|---|---|---|---|---|---|---|---|---|---|
| (unclassified single agent) | | | | | | | | | |
| Ferric carboxymaltose (unclassified single agent) | 2 | 10 vs 4 | 2.50 (0.79 –7.94) | 0.12 | 0.96 | 2.25 (0.65 –7.80) | 0.20 | 1.00; 0.80 | 0.19 / 5.23 |
| Bisphosphonate | 4 | 8 vs 3 | 2.66 (0.69 –10.20) | 0.15 | 0.96 | 2.39 (0.58 –9.91) | 0.23 | 0.61; 2.08 | 0.15 / 6.83 |
| Aldosterone antagonists (ATC C03DA) | 6 | 84 vs 67 | 1.25 (0.91 –1.72) | 0.17 | 0.96 | 1.13 (0.64 –1.97) | 0.68 | 1.15; 0.62 | 0.63 / 1.58 |
| Odanacatib (unclassified single agent) | 1 | 136 vs 115 | 1.18 (0.92 –1.51) | 0.19 | 0.96 | 1.06 (0.63 –1.79) | 0.82 | 1.26; 0.64 | 0.70 / 1.42 |
| Platelet aggregation inhibitors excl. heparin (ATC B01AC) | 4 | 88 vs 106 | 0.83 (0.63 –1.10) | 0.19 | 0.96 | 0.75 (0.43 –1.28) | 0.29 | 0.77; 1.37 | 0.67 / 1.50 |
| Lampalizumab (unclassified single agent) | 2 | 6 vs 6 | 0.50 (0.16 –1.53) | 0.22 | 0.96 | 0.45 (0.13 –1.50) | 0.19 | 1.47; — | 0.20 / 4.99 |
| Testosterone-5-alpha reductase inhibitors (ATC G04CB) | 3 | 48 vs 38 | 1.27 (0.83 –1.94) | 0.27 | 0.96 | 1.14 (0.61 –2.14) | 0.68 | 0.67; 0.77 | 0.55 / 1.83 |
| Darapladib (unclassified single agent) | 2 | 102 vs 118 | 0.86 (0.66 –1.12) | 0.27 | 0.96 | 0.78 (0.46 –1.32) | 0.35 | 1.45; 0.72 | 0.69 / 1.46 |
| PCSK9 inhibitor | 22 | 334 vs 292 | 1.09 (0.93 –1.27) | 0.29 | 0.96 | 0.98 (0.60 –1.60) | 0.93 | 1.02; 1.65* | 0.80 / 1.25 |
| Ezetimibe | 1 | 180 vs 161 | 1.12 (0.91 –1.38) | 0.29 | 0.96 | 1.01 (0.61 –1.67) | 0.98 | 1.12; 1.39 | 0.74 / 1.35 |
| Antiarrhythmics, class III (ATC C01BD) | 2 | 19 vs 26 | 0.74 (0.41 –1.33) | 0.31 | 0.96 | 0.66 (0.31 –1.40) | 0.28 | 1.68; 1.34 | 0.43 / 2.32 |
| Verubecestat (unclassified single agent) | 2 | 14 vs 4 | 1.70 (0.56 –5.11) | 0.35 | 0.96 | 1.53 (0.46 –5.05) | 0.49 | 0.36; 2.44 | 0.21 / 4.84 |
| Insulin | 6 | 6 vs 8 | 0.63 (0.23 –1.72) | 0.37 | 0.96 | 0.57 (0.19 –1.71) | 0.31 | 0.50; 2.00 | 0.24 / 4.23 |
| Omega-3 | 2 | 85 vs 74 | 1.15 (0.84 –1.56) | 0.38 | 0.96 | 1.03 (0.59 –1.80) | 0.91 | 0.80; 1.38 | 0.64 / 1.55 |
| Other blood glucose lowering drugs, | 1 | 169 vs 185 | 0.91 (0.74 –1.12) | 0.38 | 0.96 | 0.82 (0.50 –1.36) | 0.44 | 0.53*; 0.93 | 0.75 / 1.34 |

| | | | | | | | | | |
|---|---|---|---|---|---|---|---|---|---|
| excl. insulins (ATC A10BX) | | | | | | | | | |
| P2Y12 inhibitor | 4 | 220 vs 181 | 0.92 (0.75 –1.12) | 0.40 | 0.96 | 0.83 (0.50 –1.36) | 0.45 | 0.92; 1.00 | 0.75 / 1.33 |
| Alpha-adrenoreceptor antagonists (ATC G04CA) | 2 | 9 vs 13 | 0.70 (0.30 –1.63) | 0.41 | 0.96 | 0.63 (0.24 –1.65) | 0.34 | 1.01; 0.22* | 0.30 / 3.36 |
| Other antianemic preparations (ATC B03XA) | 4 | 32 vs 26 | 1.24 (0.74 –2.07) | 0.41 | 0.96 | 1.11 (0.56 –2.23) | 0.76 | 1.01; 1.21 | 0.48 / 2.09 |
| Omega-3-carboxylic acids (unclassified single agent) | 2 | 67 vs 58 | 1.15 (0.81 –1.63) | 0.43 | 0.96 | 1.03 (0.58 –1.85) | 0.91 | 1.45; 1.50 | 0.61 / 1.65 |
| Other anti-parathyroid agents (ATC H05BX) | 2 | 12 vs 16 | 0.74 (0.35 –1.57) | 0.44 | 0.96 | 0.67 (0.28 –1.61) | 0.37 | 1.79; 0.50 | 0.34 / 2.91 |
| Angiotensin II receptor blockers (ARBs), other combinations (ATC C09DX) | 6 | 56 vs 64 | 0.87 (0.61 –1.25) | 0.46 | 0.96 | 0.79 (0.44 –1.41) | 0.42 | 1.18; 0.78 | 0.60 / 1.67 |
| Other nervous system drugs (ATC N07XX) | 1 | 11 vs 8 | 1.38 (0.56 –3.40) | 0.48 | 0.96 | 1.24 (0.45 –3.41) | 0.67 | 0.00; 1.01 | 0.28 / 3.61 |
| Anti-estrogens (ATC L02BA) | 3 | 12 vs 16 | 0.77 (0.36 –1.61) | 0.48 | 0.96 | 0.69 (0.29 –1.65) | 0.40 | —; 3.07 | 0.35 / 2.89 |
| PD-1/PD-L1 (Programmed cell death protein 1/death ligand 1) inhibitors (ATC L01FF) | 49 | 8 vs 5 | 1.48 (0.50 –4.42) | 0.48 | 0.96 | 1.33 (0.41 –4.37) | 0.64 | 1.91; 1.92* | 0.21 / 4.77 |
| Other cardiac preparations (ATC C01EB) | 6 | 88 vs 95 | 0.90 (0.68 –1.20) | 0.48 | 0.96 | 0.81 (0.47 –1.40) | 0.45 | 1.23; 0.95 | 0.66 / 1.51 |
| DOAC | 11 | 64 vs 66 | 0.89 (0.63 –1.25) | 0.49 | 0.96 | 0.80 (0.45 –1.41) | 0.44 | 1.09; 0.87 | 0.61 / 1.63 |
| Investigational | 37 | 52 vs 20 | 1.18 (0.70 –2.01) | 0.53 | 0.96 | 1.06 (0.53 –2.15) | 0.86 | 2.55; 1.02 | 0.47 / 2.13 |

| | | | | | | | | | |
|---|---|---|---|---|---|---|---|---|---|
| agents without INN | | | | | | | | | |
| Omecamtiv mecarbil (unclassified single agent) | 2 | 13 vs 10 | 1.30 (0.57 –2.95) | 0.54 | 0.96 | 1.17 (0.45 –3.00) | 0.75 | 0.90; 1.02 | 0.31 / 3.24 |
| Vilanterol (unclassified single agent) | 4 | 20 vs 24 | 0.83 (0.46 –1.51) | 0.55 | 0.96 | 0.75 (0.35 –1.59) | 0.45 | 2.14; 1.23 | 0.43 / 2.33 |
| Selective beta-2-adrenoreceptor agonists (ATC R03AC) | 16 | 9 vs 5 | 1.37 (0.48 –3.89) | 0.55 | 0.96 | 1.23 (0.40 –3.85) | 0.72 | 0.71; 0.51 | 0.23 / 4.43 |
| Low-dose methotrexate | 5 | 22 vs 18 | 1.20 (0.65 –2.22) | 0.56 | 0.96 | 1.08 (0.50 –2.33) | 0.84 | —; 2.00 | 0.42 / 2.41 |
| ACE inhibitors, plain (ATC C09AA) | 1 | 7 vs 5 | 1.40 (0.45 –4.39) | 0.56 | 0.96 | 1.26 (0.37 –4.32) | 0.71 | —; — | 0.20 / 5.12 |
| Apaziquone (unclassified single agent) | 2 | 13 vs 16 | 0.81 (0.39 –1.67) | 0.57 | 0.96 | 0.73 (0.31 –1.72) | 0.47 | 0.00; 1.01 | 0.35 / 2.83 |
| SGLT2i | 46 | 348 vs 285 | 0.96 (0.82 –1.12) | 0.59 | 0.96 | 0.86 (0.53 –1.40) | 0.55 | 0.95; 1.28 | 0.80 / 1.25 |
| 3-oxoandrosten (4) derivatives (ATC G03BA) | 1 | 16 vs 19 | 0.84 (0.43 –1.64) | 0.61 | 0.96 | 0.76 (0.34 –1.70) | 0.50 | —; — | 0.39 / 2.58 |
| Sprifermin (unclassified single agent) | 1 | 31 vs 9 | 0.84 (0.41 –1.71) | 0.63 | 0.96 | 0.75 (0.32 –1.76) | 0.51 | —; — | 0.36 / 2.76 |
| GLP-1RA | 49 | 508 vs 491 | 0.97 (0.86 –1.10) | 0.63 | 0.96 | 0.87 (0.54 –1.41) | 0.58 | 0.86; 1.61* | 0.84 / 1.19 |
| Nicotinic acid and derivatives (ATC C04AC) | 1 | 25 vs 28 | 0.88 (0.52 –1.51) | 0.64 | 0.96 | 0.79 (0.39 –1.61) | 0.52 | —; 0.39 | 0.47 / 2.15 |
| TNF inhibitor | 12 | 15 vs 9 | 1.25 (0.48 –3.24) | 0.65 | 0.96 | 1.12 (0.39 –3.24) | 0.83 | 1.64; 0.29 | 0.26 / 3.90 |
| Solanezumab (unclassified single agent) | 2 | 9 vs 11 | 0.84 (0.35 –2.02) | 0.70 | 0.96 | 0.76 (0.28 –2.04) | 0.58 | 0.51; 0.00 | 0.29 / 3.50 |
| Angiotensin II receptor blockers (ARBs), plain (ATC C09CA) | 10 | 271 vs 265 | 1.03 (0.87 –1.21) | 0.75 | 0.96 | 0.92 (0.57 –1.51) | 0.75 | 1.08; 0.75 | 0.79 / 1.27 |
| Bile acids | 2 | 14 vs 6 | 1.17 (0.45 | 0.75 | 0.96 | 1.05 (0.36 | 0.93 | —; — | 0.26 / |

| | | | | | | | | | |
|---|---|---|---|---|---|---|---|---|---|
| and derivatives (ATC A05AA) | | | –3.02) | | | –3.02) | | | 3.90 |
| Carbonate (unclassified single agent) | 1 | 24 vs 26 | 0.92 (0.53 –1.58) | 0.76 | 0.96 | 0.83 (0.40 –1.69) | 0.60 | —; — | 0.46 / 2.18 |
| Other protein kinase inhibitors (ATC L01EX) | 21 | 7 vs 6 | 0.85 (0.30 –2.44) | 0.76 | 0.96 | 0.77 (0.24 –2.41) | 0.65 | 1.53; 1.73 | 0.22 / 4.49 |
| CETP inhibitor | 5 | 159 vs 152 | 1.03 (0.83 –1.29) | 0.76 | 0.96 | 0.93 (0.56 –1.55) | 0.78 | 0.97; 0.88 | 0.73 / 1.37 |
| Gantenerumab (unclassified single agent) | 2 | 7 vs 4 | 1.20 (0.33 –4.42) | 0.78 | 0.96 | 1.08 (0.27 –4.31) | 0.91 | 0.00; 0.29 | 0.16 / 6.44 |
| Other anti-dementia drugs (ATC N06DX) | 5 | 11 vs 6 | 0.87 (0.33 –2.29) | 0.78 | 0.96 | 0.78 (0.27 –2.29) | 0.66 | 0.49; 2.97 | 0.25 / 3.97 |
| Denosumab | 8 | 155 vs 149 | 1.03 (0.83 –1.29) | 0.79 | 0.96 | 0.93 (0.55 –1.55) | 0.77 | 0.72; 1.08 | 0.73 / 1.37 |
| Aspirin | 1 | 107 vs 111 | 0.96 (0.74 –1.26) | 0.79 | 0.96 | 0.87 (0.51 –1.48) | 0.60 | 1.60; 1.03 | 0.69 / 1.46 |
| CD38 (Clusters of Differentiation 38) inhibitors (ATC L01FC) | 5 | 8 vs 9 | 0.89 (0.35 –2.30) | 0.81 | 0.96 | 0.80 (0.28 –2.30) | 0.68 | 1.29; 2.40 | 0.26 / 3.87 |
| Inhaled or intranasal corticosteroids (fluticasone, mometasone; ATC D07AC) | 8 | 33 vs 27 | 1.06 (0.62 –1.80) | 0.84 | 0.96 | 0.95 (0.47 –1.92) | 0.89 | 1.17; 1.05 | 0.47 / 2.14 |
| Monoclonal antibodies (ATC L04AG) | 10 | 9 vs 4 | 1.13 (0.35 –3.64) | 0.84 | 0.96 | 1.02 (0.29 –3.58) | 0.98 | 1.19; 0.18 | 0.19 / 5.33 |
| Selective immunosuppressants (ATC L04AA) | 16 | 13 vs 8 | 0.91 (0.38 –2.21) | 0.84 | 0.96 | 0.82 (0.30 –2.23) | 0.70 | 0.40; 1.91 | 0.28 / 3.53 |
| Other vasodilators used in cardiac diseases (ATC C01DX) | 3 | 6 vs 6 | 0.89 (0.29 –2.74) | 0.84 | 0.96 | 0.80 (0.24 –2.70) | 0.72 | 1.25; 1.14 | 0.20 / 4.97 |
| Pyrimidine analogue | 1 | 12 vs 11 | 1.08 (0.48 –2.43) | 0.85 | 0.96 | 0.97 (0.38 –2.47) | 0.95 | —; — | 0.32 / 3.17 |

| | | | | | | | | | |
|---|---|---|---|---|---|---|---|---|---|
| s (ATC L01BC) | | | | | | | | | |
| Creatine (unclassified single agent) | 1 | 26 vs 27 | 0.96 (0.56 –1.62) | 0.87 | 0.96 | 0.86 (0.43 –1.74) | 0.67 | —; 0.99 | 0.47 / 2.13 |
| Poly (ADP-ribose) polymerase (PARP) inhibitors (ATC L01XK) | 10 | 7 vs 5 | 1.11 (0.32 –3.91) | 0.87 | 0.96 | 1.00 (0.26 –3.82) | 1.00 | 0.48; 1.00 | 0.17 / 6.03 |
| Statin | 4 | 19 vs 20 | 0.95 (0.51 –1.78) | 0.87 | 0.96 | 0.85 (0.39 –1.86) | 0.69 | 1.00; 1.21 | 0.41 / 2.45 |
| Calcitonin | 4 | 47 vs 44 | 1.03 (0.69 –1.56) | 0.87 | 0.96 | 0.93 (0.50 –1.73) | 0.82 | 0.63; — | 0.56 / 1.80 |
| Bempedoic acid | 3 | 76 vs 74 | 0.98 (0.71 –1.34) | 0.89 | 0.96 | 0.88 (0.50 –1.54) | 0.65 | 1.02; 1.19 | 0.64 / 1.57 |
| Centrally acting antiobesity products (ATC A08AA) | 9 | 244 vs 245 | 0.99 (0.83 –1.18) | 0.90 | 0.96 | 0.89 (0.54 –1.46) | 0.64 | 1.05; 0.58 | 0.78 / 1.29 |
| Thiazolidinedione | 7 | 25 vs 24 | 0.97 (0.56 –1.70) | 0.92 | 0.97 | 0.87 (0.42 –1.80) | 0.72 | 0.86; 1.75 | 0.45 / 2.22 |
| JAK inhibitor | 17 | 11 vs 6 | 1.02 (0.38 –2.69) | 0.98 | 1.00 | 0.91 (0.31 –2.68) | 0.87 | 0.50; — | 0.25 / 4.02 |
| Fibrate | 2 | 39 vs 39 | 1.00 (0.64 –1.56) | 0.99 | 1.00 | 0.90 (0.48 –1.71) | 0.75 | 1.31; 1.84 | 0.53 / 1.88 |
| Intravitreal or systemic corticosteroids (triamcinolone, dexamethasone; ATC A01AC) | 4 | 8 vs 4 | 1.00 (0.30 –3.32) | 1.00 | 1.00 | 0.90 (0.25 –3.25) | 0.87 | 3.06*; 0.25 | 0.18 / 5.53 |
| Apolipoprotein a-i (unclassified single agent) | 1 | 5 vs 5 | 1.00 (0.29 –3.46) | 1.00 | 1.00 | 0.90 (0.24 –3.38) | 0.88 | —; — | 0.17 / 5.88 |

Negative-control columns flag (*) class-level risk ratios for cataract or inguinal-hernia SAEs with nominal $P < 0.05$ (protocol section 8). Detectable RR: the smallest risk ratio below and above 1 that the class estimate could detect with 80% power at two-sided $\alpha = 0.05$, given its standard error. ATC-derived classes take the first ATC code returned by RxNav and can group agents given by different routes: ATC D07AC contains inhaled and intranasal fluticasone and mometasone (respiratory trials); ATC A01AC contains intravitreal triamcinolone and systemic dexamethasone. Calibrated estimates use the empirical null distribution fitted to all class-level negative-control-outcome estimates with at least 10 events.

### Supplementary Table S3. Meta-regression of trial log risk ratio on adverse-event reporting window, and competing-mortality sensitivity

**(a) Meta-regression** (inverse-variance weighted; classes with ≥5 informative trials with a stated window; shown for pre-specified classes and any class with P < 0.05; not adjusted for multiplicity)

| Class | Trials | Slope (log RR per year) | SE | P |
|---|---|---|---|---|
| Anti-NGF | 6 | +0.309 | 0.381 | 0.463 |
| DOAC | 5 | -0.537 | 0.410 | 0.281 |
| DPP-4i | 11 | +0.141 | 0.079 | 0.106 |
| GLP-1RA | 38 | +0.069 | 0.031 | 0.0338 |
| PCSK9 inhibitor | 22 | +0.104 | 0.043 | 0.0241 |
| SGLT2i | 35 | +0.038 | 0.036 | 0.301 |

**(b) Competing mortality** (at-risk counts rescaled to N × (1 − ½ × proportion dying) in each arm)

| Class | Primary RR (95% CI) | Survivor-adjusted RR (95% CI) |
|---|---|---|
| GLP-1RA | 0.97 (0.86-1.10) | 0.97 (0.85-1.09) |
| SGLT2i | 0.96 (0.82-1.12) | 0.95 (0.82-1.12) |
| DPP-4i | 1.26 (0.99-1.61) | 1.26 (0.99-1.61) |
| IL-1 inhibitor | 0.57 (0.39-0.82) | 0.57 (0.39-0.82) |
| Anti-NGF | 2.31 (1.82-2.92) | 2.31 (1.82-2.91) |
| Statin | 0.95 (0.51-1.78) | 0.95 (0.51-1.78) |
| PCSK9 inhibitor | 1.09 (0.93-1.27) | 1.09 (0.93-1.27) |

### Supplementary Table S4. Within-trial comparison of the highest vs lowest randomized dose (classes with ≥10 events)

| Class | Trials | Events (high vs low) | RR high vs low dose (95% CI) | P |
|---|---|---|---|---|
| Investigational agents without INN | 23 | 12 vs 5 | 2.22 (0.83–5.98) | 0.11 |
| DOAC | 7 | 26 vs 39 | 0.69 (0.42–1.13) | 0.14 |
| GLP-1RA | 36 | 22 vs 29 | 0.67 (0.39–1.17) | 0.16 |
| SGLT2i | 25 | 62 vs 76 | 0.80 (0.58–1.12) | 0.20 |
| Anti-NGF | 13 | 58 vs 126 | 1.24 (0.87–1.78) | 0.23 |
| Otilimab (unclassified single agent) | 2 | 4 vs 6 | 0.67 (0.19–2.38) | 0.54 |
| Verubecestat (unclassified single agent) | 2 | 4 vs 7 | 0.76 (0.25–2.34) | 0.63 |
| JAK inhibitor | 21 | 58 vs 60 | 0.93 (0.65–1.33) | 0.70 |
| IL-1 inhibitor | 2 | 19 vs 18 | 1.01 (0.53–1.92) | 0.97 |
| P2Y12 inhibitor | 1 | 54 vs 54 | 1.00 (0.68–1.45) | 0.98 |
| Intravitreal or systemic corticosteroids (triamcinolone, dexamethasone; ATC A01AC) | 4 | 5 vs 5 | 0.99 (0.29–3.41) | 0.98 |

Dose was read from the reporting-group title or description as the mass or unit value that follows the name of the drug under test (correction C8); one contrast per trial and drug; primary-window trials only. Comparisons are randomized within trial.

### Supplementary Table S5. Pre-specified estimates before and after four rounds of error correction

#### (a) Correction of the adverse-event window parser after the first analysis (deviation D4)

| Analysis | Before correction: trials | Before: RR (95% CI) | After correction: trials | After: RR (95% CI) |
|---|---|---|---|---|
| P1 Canakinumab → OA-related SAE | 3 | 0.57 (0.39–0.82) | 3 | 0.57 (0.39–0.82) |
| P2 Anti-NGF → OA-related SAE | 14 | 2.34 (1.85–2.95) | 14 | 2.34 (1.85–2.95) |
| P3 SGLT2i → gout | 42 | 0.66 (0.45–0.99) | 45 | 0.66 (0.45–0.99) |
| H1 GLP-1RA | 48 | 0.94 (0.82–1.07) | 51 | 0.96 (0.85–1.09) |
| H1b GLP-1RA, outside the preliminary analysis | 36 | 0.97 (0.58–1.64) | 38 | 0.98 (0.59–1.63) |
| H2 SGLT2i | 42 | 0.95 (0.81–1.11) | 45 | 0.95 (0.81–1.11) |
| H3 DPP-4i | 26 | 1.24 (0.97–1.58) | 29 | 1.22 (0.96–1.55) |
| H3b DPP-4i, outside the preliminary analysis | 21 | 0.92 (0.39–2.14) | 24 | 0.85 (0.41–1.74) |
| H4 IL-1β inhibition | 4 | 0.58 (0.40–0.83) | 3 | 0.57 (0.39–0.82) |
| H5 Anti-NGF | 14 | 2.34 (1.85–2.95) | 14 | 2.34 (1.85–2.95) |
| H6 Bone-active agents | 12 | 0.98 (0.79–1.22) | 13 | 1.00 (0.81–1.24) |
| H7 Statins + PCSK9i | 26 | 1.00 (0.84–1.20) | 28 | 1.08 (0.93–1.26) |
| H8 VKA vs DOAC (head-to-head) | 15 | 1.11 (0.93–1.33) | 15 | 1.11 (0.93–1.33) |

#### (b) Corrections C1–C9 after internal review (Protocol addendum D7: window offsets, VKA comparators, class mapping, nested and contaminated groups, missing denominators)

| Analysis | Before D7: trials | Before D7: RR (95% CI) | After D7: trials | After D7: RR (95% CI) |
|---|---|---|---|---|
| P1 Canakinumab → OA-related SAE | 3 | 0.57 (0.39–0.82) | 3 | 0.57 (0.39–0.82) |
| P2 Anti-NGF → OA-related SAE | 14 | 2.34 (1.85–2.95) | 14 | 2.34 (1.85–2.95) |
| P3 SGLT2i → gout | 45 | 0.66 (0.45–0.99) | 45 | 0.66 (0.45–0.99) |
| H1 GLP-1RA | 51 | 0.96 (0.85–1.09) | 51 | 0.97 (0.86–1.10) |
| H1b GLP-1RA, outside the preliminary analysis | 38 | 0.98 (0.59–1.63) | 38 | 1.13 (0.68–1.88) |
| H2 SGLT2i | 45 | 0.95 (0.81–1.11) | 45 | 0.96 (0.82–1.12) |
| H3 DPP-4i | 29 | 1.22 (0.96–1.55) | 31 | 1.22 (0.96–1.56) |
| H3b DPP-4i, outside the preliminary analysis | 24 | 0.85 (0.41–1.74) | 26 | 0.90 (0.44–1.83) |
| H4 IL-1β inhibition | 3 | 0.57 (0.39–0.82) | 3 | 0.57 (0.39–0.82) |
| H5 Anti-NGF | 14 | 2.34 (1.85–2.95) | 14 | 2.34 (1.85–2.95) |
| H6 Bone-active agents | 13 | 1.00 (0.81–1.24) | 13 | 1.00 (0.81–1.24) |
| H7 Statins + PCSK9i | 28 | 1.08 (0.93–1.26) | 27 | 1.08 (0.93–1.26) |
| H8 VKA vs DOAC (head-to-head) | 15 | 1.11 (0.93–1.33) | 20 | 1.12 (0.95–1.33) |

#### (c) Corrections C10–C11 after simulated peer review (Protocol addendum D9: combination arms counted twice within a class; on-treatment adverse-event windows read as 0–1 week, set to not stated after reading all 87 large or informative trials with windows under 24 weeks)

| Analysis | Before D9: trials | Before D9: RR (95% CI) | After D9: trials | After D9: RR (95% CI) |
|---|---|---|---|---|
| P1 Canakinumab → OA-related SAE | 3 | 0.57 (0.39–0.82) | 3 | 0.57 (0.39–0.82) |
| P2 Anti-NGF → OA-related SAE | 14 | 2.34 (1.85–2.95) | 14 | 2.34 (1.85–2.95) |
| P3 SGLT2i → gout | 45 | 0.66 (0.45–0.99) | 46 | 0.66 (0.45–0.99) |
| H1 GLP-1RA | 51 | 0.97 (0.86–1.10) | 51 | 0.97 (0.86–1.10) |
| H1b GLP-1RA, outside the preliminary analysis | 38 | 1.13 (0.68–1.88) | 38 | 1.13 (0.68–1.88) |
| H2 SGLT2i | 45 | 0.96 (0.82–1.12) | 46 | 0.96 (0.82–1.12) |
| H3 DPP-4i | 31 | 1.22 (0.96–1.56) | 32 | 1.22 (0.96–1.56) |
| H3b DPP-4i, outside the preliminary analysis | 26 | 0.90 (0.44–1.83) | 27 | 0.90 (0.44–1.83) |
| H4 IL-1β inhibition | 3 | 0.57 (0.39–0.82) | 3 | 0.57 (0.39–0.82) |
| H5 Anti-NGF | 14 | 2.34 (1.85–2.95) | 14 | 2.34 (1.85–2.95) |
| H6 Bone-active agents | 13 | 1.00 (0.81–1.24) | 13 | 1.00 (0.81–1.24) |
| H7 Statins + PCSK9i | 27 | 1.08 (0.93–1.26) | 27 | 1.08 (0.93–1.26) |
| H8 VKA vs DOAC (head-to-head) | 20 | 1.12 (0.95–1.33) | 22 | 1.12 (0.94–1.32) |
| H8b VKA vs DOAC, all windows | 23 | 1.12 (0.94–1.32) | 23 | 1.12 (0.94–1.32) |

#### (d) Corrections C12–C18 after an automated re-audit of the verification workbook (Protocol addendum D10: nested and extension records, groups mixing randomized arms or reported over different periods, active or strategy comparators, wrong comparator, wrongly excluded trials, windows read from truncated text), with screens S1–S4 of all included trials

| Analysis | Before D10: trials | Before D10: RR (95% CI) | After D10: trials | After D10: RR (95% CI) |
|---|---|---|---|---|
| P1 Canakinumab → OA-related SAE | 3 | 0.57 (0.39–0.82) | 3 | 0.57 (0.39–0.82) |
| P2 Anti-NGF → OA-related SAE | 14 | 2.34 (1.85–2.95) | 12 | 2.31 (1.82–2.92) |
| P3 SGLT2i → gout | 46 | 0.66 (0.45–0.99) | 46 | 0.66 (0.45–0.99) |
| H1 GLP-1RA | 51 | 0.97 (0.86–1.10) | 49 | 0.97 (0.86–1.10) |
| H1b GLP-1RA, outside the preliminary analysis | 38 | 1.13 (0.68–1.88) | 36 | 1.12 (0.67–1.88) |
| H2 SGLT2i | 46 | 0.96 (0.82–1.12) | 46 | 0.96 (0.82–1.12) |
| H3 DPP-4i | 32 | 1.22 (0.96–1.56) | 30 | 1.26 (0.99–1.61) |
| H3b DPP-4i, outside the preliminary analysis | 27 | 0.90 (0.44–1.83) | 25 | 1.12 (0.49–2.53) |
| H4 IL-1β inhibition | 3 | 0.57 (0.39–0.82) | 3 | 0.57 (0.39–0.82) |
| H5 Anti-NGF | 14 | 2.34 (1.85–2.95) | 12 | 2.31 (1.82–2.92) |
| H6 Bone-active agents | 13 | 1.00 (0.81–1.24) | 13 | 1.00 (0.81–1.24) |
| H7 Statins + PCSK9i | 27 | 1.08 (0.93–1.26) | 26 | 1.08 (0.93–1.26) |
| H8 VKA vs DOAC (head-to-head) | 22 | 1.12 (0.94–1.32) | 21 | 1.10 (0.92–1.30) |
| H8b VKA vs DOAC, all | 23 | 1.12 (0.94–1.32) | 22 | 1.10 (0.92–1.30) |

| windows |
|---|

Screens S1–S4 listed 1,120 hits in 732 trials, each read against the registry text by language-model reviewers without outcome counts (a consistency re-check of every kept switch-related hit; analyst decisions on unresolved rows; 2 errors found incidentally; decisions in `adjudication/audit_D10.tsv`). With section 1 of the addendum, D10 excluded 14 trials, dropped 86 comparisons in 69 trials and re-specified or restored the comparisons of 23 trials. The primary set lost 74 trials (51 whose placebo or usual-care comparisons were dropped; 12 excluded; 11 now head-to-head) and gained 2.

**(e) VKA vs DOAC without trials that assign different antiplatelet co-therapy by arm (Protocol addendum D10, R8; PIONEER AF-PCI, RE-DUAL PCI, ENTRUST-AF PCI, PETRO)**

| Analysis | Trials | Events (VKA vs DOAC) | RR (95% CI) |
|---|---|---|---|
| H8 VKA vs DOAC: OA-related SAE, all trials | 21 | 240 vs 311 | 1.10 (0.92–1.30) |
| H8 VKA vs DOAC: OA-related SAE, same antiplatelet background | 18 | 233 vs 293 | 1.12 (0.94–1.34) |
| H8b VKA vs DOAC: OA-related SAE, all trials | 22 | 240 vs 312 | 1.10 (0.92–1.30) |
| H8b VKA vs DOAC: OA-related SAE, same antiplatelet background | 18 | 233 vs 293 | 1.12 (0.94–1.34) |
| M4 DOAC vs VKA: intracranial haemorrhage, all trials | 21 | 419 vs 480 | 0.64 (0.56–0.73) |
| M4 DOAC vs VKA: intracranial haemorrhage, same antiplatelet background | 18 | 410 vs 463 | 0.65 (0.58–0.75) |

Intracranial haemorrhage is shown as DOAC vs VKA. All results in the main text use the corrected analysis (after D10). Earlier results are archived with the analysis code.

### Supplementary Table S6. Published observational estimates of hip or knee replacement and randomized atlas estimates (systematic search; Protocol addendum D9, R6–R7)

**(a) Estimates compared** (21 counted, from 16 studies; 8 differ at P < 0.05, from 5 studies; 9 with the sensitivity comparators; 3 and 6 after calibration with the primary and refitted nulls; 6 compatible and informative; 7 uninformative)

| Study (PMID) | Contrast | Setting · joint | Observational (95% CI) | Randomized (95% CI) | Ratio (95% CI) | P | P calibrated (primary; refitted) | Class | Randomized comparator |
|---|---|---|---|---|---|---|---|---|---|
| Porto 2025 | GLP-1RA vs non- | obese, no OA · hip | HR 1.13 (0.69– | 1.04 (0.80 –1.36) | 1.09 (0.62 –1.90) | 0.77 | 0.61; 0.67 | compatible and | GLP-1RA (obesity |

| | | | | | | | | | |
|---|---|---|---|---|---|---|---|---|---|
| (39811151) | use | | 1.85) | | | | | informative | trials) |
| Porto 2025 (39811151) | GLP-1RA vs non-use | obese, no OA · knee | HR 2.61 (1.60–4.25) | 1.04 (0.80–1.36) | 2.51 (1.44–4.37) | 0.001 | 0.005; 0.002 | differs | GLP-1RA (obesity trials) |
| Porto 2025 (39811151) | GLP-1RA vs non-use | obese, hip OA · hip | HR 0.60 (0.45–0.79) | 1.04 (0.80–1.36) | 0.58 (0.39–0.85) | 0.005 | 0.15; 0.033 | differs | GLP-1RA (obesity trials) |
| Porto 2025 (39811151) | GLP-1RA vs non-use | obese, knee OA · knee | HR 0.75 (0.63–0.89) | 1.04 (0.80–1.36) | 0.72 (0.53–0.99) | 0.043 | 0.44; 0.18 | differs | GLP-1RA (obesity trials) |
| Lin 2025 (39928811) | GLP-1RA vs non-use | T2D, knee OA · knee | HR 0.91 (0.89–0.98) | 0.95 (0.83–1.09) | 0.96 (0.83–1.11) | 0.59 | 0.79; 0.95 | compatible and informative | GLP-1RA (T2D/other trials) |
| Carter 2026 (42229941) | GLP-1RA vs non-use | knee OA · knee | HR 0.90 (0.83–0.98) | 0.97 (0.86–1.10) | 0.93 (0.80–1.08) | 0.32 | 0.90; 0.87 | compatible and informative | GLP-1RA |
| Su 2025 (40478380) | SGLT2i vs GLP-1RA | T2D · hip or knee | HR 0.70 (0.55–0.90) | 1.12 (0.88–1.42) † | 0.63 (0.45–0.89) | 0.008 | 0.22; 0.064 | differs | SGLT2i (without heart-failure and kidney trials) / GLP-1RA (T2D/other trials) |
| Jao 2026 (42475344) | SGLT2i vs GLP-1RA | T2D · hip | HR 1.17 (0.86–1.58) | 1.12 (0.88–1.42) † | 1.05 (0.71–1.54) | 0.82 | 0.91; 0.87 | compatible and informative | SGLT2i (without heart-failure and kidney trials) / GLP-1RA (T2D/other trials) |
| Jao 2026 (42475344) | SGLT2i vs GLP-1RA | T2D · knee | HR 0.88 (0.65–1.18) | 1.12 (0.88–1.42) † | 0.79 (0.54–1.15) | 0.22 | 0.53; 0.36 | compatible and informative | SGLT2i (without heart-failure and kidney trials) / GLP-1RA (T2D/other trials) |
| Liu 2026 (42139974) | SGLT2i vs GLP-1RA | T2D · any joint | HR 0.85 (0.73–1.00) | 1.12 (0.88–1.42) † | 0.76 (0.57–1.01) | 0.063 | 0.45; 0.24 | compatible and informative | SGLT2i (without heart-failure and kidney trials) / GLP-1RA (T2D/other trials) |
| Hu 2026 (41886946) | SGLT2i vs non-use | T2D · knee | HR 1.33 (1.25–1.42) | 0.96 (0.82–1.12) | 1.39 (1.17–1.65) | 1.6e-04 | 0.084; 0.013 | differs | SGLT2i |
| Chen 2026 (41919008) | DPP-4i vs non-use | T2D · hip | HR 0.38 (0.30–0.48) | 1.26 (0.99–1.61) | 0.30 (0.21–0.42) | 4.1e-12 | 1.9e-04; 7.8e-08 | differs | DPP-4i |

| | | | | | | | | | |
|---|---|---|---|---|---|---|---|---|---|
| Chen 2026 (41919008) | DPP-4i vs non-use | T2D · knee | HR 0.42 (0.37–0.46) | 1.26 (0.99–1.61) | 0.33 (0.26–0.44) | 7.9e-16 | 2.7e-04; 1.6e-08 | differs | DPP-4i |
| Liu 2026 (42139974) | SGLT2i vs DPP-4i | T2D · any joint (direction unclear) | HR 1.10 (0.95–1.27) | 0.84 (0.62–1.15) † | 1.31 (0.92–1.84) | 0.13 | 0.48; 0.29 | not counted (direction unclear) | SGLT2i (without heart-failure and kidney trials) / DPP-4i |
| Liu 2026 (42139974) | GLP-1RA vs DPP-4i | T2D · any joint (direction unclear) | HR 1.22 (1.07–1.39) | 0.75 (0.57–1.00) † | 1.62 (1.19–2.20) | 0.002 | 0.19; 0.044 | not counted (direction unclear) | GLP-1RA (T2D/other trials) / DPP-4i |
| Fu 2017 (28590379) | Bisphosphonate vs non-use | osteoporosis, knee OA · knee | HR 0.76 (0.69–0.83) | 2.66 (0.69–10.20) | 0.29 (0.07–1.10) | 0.068 | 0.11; 0.085 | uninformative | Bisphosphonate |
| Hung 2018 (30306268) | Bisphosphonate vs non-use | hip OA · hip | HR 0.97 (0.74–1.27) | 2.66 (0.69–10.20) | 0.37 (0.09–1.44) | 0.15 | 0.22; 0.18 | uninformative | Bisphosphonate |
| Neogi 2018 (28988204) | Bisphosphonate vs non-use | women, knee OA · knee | HR 0.76 (0.60–0.95) | 2.66 (0.69–10.20) | 0.29 (0.07–1.12) | 0.072 | 0.12; 0.089 | uninformative | Bisphosphonate |
| Wang J 2026 (42750457) | Bisphosphonate vs non-use | UK Biobank · knee | HR 0.91 (0.67–1.24) | 2.66 (0.69–10.20) | 0.34 (0.09–1.36) | 0.13 | 0.19; 0.15 | uninformative | Bisphosphonate |
| Yu 2024 (39768410) | Denosumab vs bisphosphonate | women, knee OA · knee | HR 0.77 (0.62–0.97) | 0.39 (0.10–1.51) † | 1.99 (0.50–7.91) | 0.33 | 0.38; 0.34 | uninformative | Denosumab / Bisphosphonate |
| Chen JY 2025 (41157204) | Denosumab vs bisphosphonate | women, post-traumatic knee OA · knee | HR 1.09 (0.88–1.35) | 0.39 (0.10–1.51) † | 2.81 (0.71–11.17) | 0.14 | 0.18; 0.15 | uninformative | Denosumab / Bisphosphonate |
| Sarmanova 2020 (32097491) | Statin vs non-use | general population · hip or knee | HR 0.99 (0.97–1.03) | 0.95 (0.51–1.78) | 1.04 (0.56–1.96) | 0.89 | 0.71; 0.79 | uninformative | Statin |
| Ballal 2021 (34380108) | Warfarin vs DOAC | atrial fibrillation · hip or knee | OR 1.59 (1.31–1.92) | 1.10 (0.92–1.30) | 1.45 (1.12–1.87) | 0.005 | 0.077; 0.020 | differs | VKA vs DOAC |

### (b) Eligible estimates not counted

| Study (PMID) | Exposure | Comparator | Outcome | Estimate (95% CI) | Reason |
|---|---|---|---|---|---|
| Lavu 2024 (38857711) | GLP-1RA prescribed | GLP-1RA not prescribed (PS-matched) | THA; TKA (separate outcomes) | — | estimate not extractable (hazard ratios only in closed full text) |
| Carter 2026 (42229941) | Semaglutide or tirzepatide ('new generation'), 3-year exposure | PS-matched non-exposed | TKA (cumulative incidence) | HR 0.72 (0.67–0.78) | secondary exposure definition (semaglutide or tirzepatide, 3 years) |
| Wang Z 2026 (41235723) | Colchicine initiators (oral) | PS-matched colchicine non-initiators | TJA (TKA or THA) | HR 0.89 (0.82–0.97) | no randomized comparator (no colchicine trial contributed events) |
| Wang Z 2026 | Colchicine | PS-matched | TKA | HR 0.94 (0.85– | joint-specific |

| | | | | | |
|---|---|---|---|---|---|
| (41235723) | initiators (oral) | colchicine non-initiators | | 1.05) | sensitivity analysis |
| Wang Z 2026 (41235723) | Colchicine initiators (oral) | PS-matched colchicine non-initiators | THA | HR 0.87 (0.77–0.98) | joint-specific sensitivity analysis |
| Wang Z 2026 (41235723) | Colchicine initiators (oral) | PS-matched colchicine non-initiators | TJA (TKA or THA) | HR 0.76 (0.63–0.91) | population subgroup |
| Wang Z 2026 (41235723) | Colchicine initiators (oral) | PS-matched colchicine non-initiators | TJA (TKA or THA) | HR 0.89 (0.79–1.01) | population subgroup |
| Wang Z 2026 (41235723) | Colchicine initiators (oral) | PS-matched colchicine non-initiators | TJA (TKA or THA) | HR 0.93 (0.83–1.05) | population subgroup |
| Sarmanova 2020 (32097491) | Statin use (any) | Never prescribed statins (matched on age, gender, practice, PS) | Knee or hip replacement due to OA | HR 0.97 (0.94–1.01) | outcome by indication (secondary) |
| Sarmanova 2020 (32097491) | Statin use (any) | Never prescribed statins (matched on age, gender, practice, PS) | Knee or hip replacement due to RA | HR 0.77 (0.63–0.94) | outcome by indication (secondary) |
| Sarmanova 2020 (32097491) | High-intensity statin (LDL reduction 42-55%) | Never prescribed statins (matched on age, gender, practice, PS) | Knee or hip joint replacement (overall) | HR 0.86 (0.75–0.98) | exposure-intensity subgroup |
| Chen JY 2025 (41157204) | Raloxifene (not a listed class) | Bisphosphonates (reference) | TKR | HR 0.81 (0.67–0.99) | exposure (raloxifene) not a pre-specified class |
| Ballal 2021 (34380108) | Warfarin (VKA) use within 1 y before index | DOAC use within 1 y before index | Knee replacement | OR 1.58 (1.22–2.04) | joint-specific sensitivity analysis |
| Ballal 2021 (34380108) | Warfarin (VKA) use within 1 y before index | DOAC use within 1 y before index | Hip replacement | OR 1.33 (1.03–1.72) | joint-specific sensitivity analysis |

Search: PubMed and Europe PMC, inception to 11 October 2026, eligibility fixed before searching (Supplementary File 6); 7,282 hits, 3,487 unique records screened, 26 full texts or abstracts assessed, 18 studies eligible (search strings, screening decisions and flow in the repository, `obs_search_D9/`). For each study the authors' primary estimate per comparison, population and joint was used; secondary exposure definitions, indications and sensitivity analyses are listed in (b). Randomized comparators follow the addendum: GLP-1RA trials by population (obesity; diabetes and other; all), bisphosphonate trials for bisphosphonates, and SGLT2 inhibitor trials outside heart-failure and kidney populations in indirect comparisons (Bucher method, †). The P value tests the difference between log estimates; class: differs ($P < 0.05$), compatible and informative ($P \geq 0.05$ and randomized upper/lower CI ratio ≤ 2), uninformative (otherwise). Liu 2026 reports hazard ratios above 1 together with lower absolute risks in the first-named group for both contrasts with DPP-4 inhibitors; oriented as those risks imply, both are compatible (8 of 23 would differ). Calibrated P values widen the randomized interval for systematic error with the empirical null (variance + $SD^2$, shifted by the null mean for direct contrasts; + 2 $SD^2$ without shift for indirect contrasts). The earlier targeted search (addendum D7, 11 estimates from 7 studies) is superseded; all its estimates are included here, and its comparison is in Supplementary Data.

### Supplementary Table S7. Estimates inside and outside the preliminary analysis (post hoc, addendum A1)

| Analysis | Share of events previewed | Previewed trials: RR (95% CI) | Trials | Outside the look: RR (95% CI) |
|---|---|---|---|---|

The preliminary pooled analysis of 72 large trials (Supplementary File 3) preceded the protocol. Trials: number of trials outside the look.

### Supplementary Table S8. The OA-related SAE construct (post hoc, addendum A2)

#### (a) Pre-specified analyses by term type

| Analysis | All OA-related terms | Arthroplasty or replacement terms only | Osteoarthritis terms only | Maximum-term (participant) counting |
|---|---|---|---|---|
| P1 canakinumab | 0.57 (0.39–0.82) | — | 0.57 (0.39–0.82) | 0.57 (0.39–0.82) |
| P2/H5 anti-NGF | 2.31 (1.82–2.92) | 1.06 (0.73–1.52) | 1.71 (1.15–2.53) | 3.08 (2.19–4.34) |
| H1 GLP-1RA | 0.97 (0.86–1.10) | 0.66 (0.26–1.66) | 0.98 (0.86–1.11) | 0.98 (0.86–1.11) |
| H2 SGLT2i | 0.96 (0.82–1.12) | 0.75 (0.37–1.53) | 0.97 (0.82–1.14) | 0.99 (0.84–1.16) |
| H3 DPP-4i | 1.26 (0.99–1.61) | — | 1.26 (0.99–1.61) | 1.26 (0.99–1.61) |
| H4 IL-1 inhibition | 0.57 (0.39–0.82) | — | 0.57 (0.39–0.82) | 0.57 (0.39–0.82) |
| H6 bone-active agents | 1.00 (0.81–1.24) | 4.94 (0.58–42.33) | 0.98 (0.79–1.21) | 0.98 (0.79–1.21) |
| H7 statin + PCSK9i | 1.08 (0.93–1.26) | 2.33 (0.60–9.04) | 1.07 (0.92–1.25) | 1.08 (0.92–1.25) |
| H8 VKA vs DOAC | 1.10 (0.92–1.30) | 0.98 (0.48–1.99) | 1.10 (0.93–1.32) | 1.10 (0.92–1.31) |

#### (b) Anti-NGF antibodies (P2 analysis set)

| Outcome | Trials | Events (A vs C) | RR (95% CI) |
|---|---|---|---|
| OA-related SAE (P2) | 12 | 584 vs 77 | 2.31 (1.82–2.92) |
| rapidly progressive OA / chondrolysis terms | 12 | 302 vs 11 | 8.07 (4.41–14.77) |
| osteoarthritis terms | 12 | 144 vs 30 | 1.71 (1.15–2.53) |
| arthroplasty terms | 12 | 138 vs 36 | 1.06 (0.73–1.52) |
| joint-safety composite (+ osteonecrosis, subchondral insufficiency fracture) | 12 | 645 vs 81 | 2.47 (1.97–3.10) |
| OA-related SAE, clinically used doses | 11 | 272 vs 77 | 1.73 (1.34–2.22) |
| arthroplasty terms, clinically used doses | 11 | 90 vs 36 | 1.10 (0.75–1.63) |

Rapidly progressive OA and chondrolysis are terms used almost only in anti-NGF programmes. Composite: OA-related terms plus osteonecrosis (not of the jaw) and subchondral insufficiency fracture. Clinically used doses: tanezumab ≤5 mg, fasinumab ≤1 mg, fulranumab ≤3 mg.

### Supplementary Table S9. Modest-effect positive controls (addendum A6; expected effects fixed before extraction)

| Positive control | Trials | Events (A vs C) | RR (95% CI) | Expected (published, 95% CI) | Recovered |
|---|---|---|---|---|---|
| M1 GLP-1RA → gallbladder/biliary | 49 | 862 vs 548 | 1.36 (1.22–1.51) | 1.37 (1.23–1.52) | yes |
| M2 SGLT2i → ketoacidosis | 46 | 188 vs 72 | 1.96 (1.48–2.60) | 2.13 (1.38–3.27) | yes |
| M3 thiazolidinedione → heart failure | 7 | 21 vs 10 | 1.42 (0.63–3.18) | 1.72 (1.21–2.42) | no |
| M4 DOAC vs VKA → intracranial haemorrhage | 21 | 419 vs 480 | 0.64 (0.56–0.73) | 0.48 (0.39–0.59) | yes |
| M5 SGLT2i → heart failure | 46 | 3326 vs 4209 | 0.74 (0.71–0.77) | 0.69 (0.61–0.79) | yes |

Recovered: 95% CI excluding 1 in the expected direction. Sources of the expected effects: He et al. 2022 (M1), Liu et al. 2020 (M2, odds ratio), Lago et al. 2007 (M3), Ruff et al. 2014 (M4), Zelniker et al. 2019 (M5, hazard ratio). M4 is shown as DOAC vs VKA. Analysis sets as for the hypotheses, within the 1,699 adjudicated trials.

### Supplementary Table S10. Empirical calibration (addendum A3)

#### (a) Null distributions and classes significant after calibration

| Null fitted to | Estimates | Mean (SD), log scale | Significant after calibration | Calibrated P: anti-NGF; IL-1β |
|---|---|---|---|---|
| All negative-control estimates (primary) | 100 | 0.106 (0.236) | Anti-androgens (P = 0.028), Anti-NGF, IL-1β inhibition | 0.0058; 0.0269 |
| Without GLP-1RA × inguinal hernia | 99 | 0.098 (0.232) | Anti-androgens (P = 0.027), Anti-NGF, IL-1β inhibition | 0.0047; 0.0271 |
| Without estimates with a known cause (A3) | 96 | 0.051 (0.126) | Anti-androgens (P = 0.012), Anti-NGF, IL-1β inhibition | <0.0001; 0.0068 |
| Cataract only | 53 | 0.140 (0.262) | Anti-androgens (P = 0.038), Anti-NGF, IL-1β inhibition | 0.0156; 0.0294 |
| Inguinal hernia only | 47 | 0.069 (0.196) | Anti-androgens (P = 0.019), Anti-NGF, IL-1β inhibition | 0.0008; 0.0201 |

GLP-1RAs × inguinal hernia (RR 1.61): calibrated P = 0.16 against the primary null and 0.020 against the null without known-effect estimates, each fitted without the estimate itself. PCSK9 inhibitors × inguinal hernia (RR 1.65): calibrated P = 0.16 against the primary null and 0.013 against the null without known-effect estimates, each fitted without the estimate itself.

#### (b) Calibrated estimates for the pre-specified analyses

| Analysis | Calibrated RR, null without known-effect estimates | Calibrated RR, primary null |
|---|---|---|

| | | |
|---|---|---|
| P1 canakinumab | 0.54 (0.35–0.84) | 0.51 (0.28–0.93) |
| P2/H5 anti-NGF | 2.19 (1.56–3.08) | 2.07 (1.23–3.48) |
| H1 GLP-1RA | 0.92 (0.70–1.21) | 0.87 (0.54–1.41) |
| H2 SGLT2i | 0.91 (0.68–1.22) | 0.86 (0.53–1.40) |
| H3 DPP-4i | 1.20 (0.85–1.69) | 1.13 (0.67–1.91) |
| H4 IL-1 inhibition | 0.54 (0.35–0.84) | 0.51 (0.28–0.93) |
| H6 bone-active agents | 0.95 (0.69–1.32) | 0.90 (0.54–1.50) |
| H7 statin + PCSK9i | 1.03 (0.77–1.37) | 0.97 (0.60–1.58) |
| H8 VKA vs DOAC | 1.04 (0.77–1.41) | 0.99 (0.60–1.61) |

Known-effect negative-control estimates: corticosteroids (any route) with cataract, GLP-1RA with inguinal hernia, and any class–outcome pair with nominal P < 0.001.

### Supplementary Table S11. Precision of the pre-specified analyses and GLP-1RA by molecule (addenda A4, A5)

#### (a) Smallest risk ratio detectable with 80% power (two-sided α = 0.05) given the observed standard error

| Analysis | Detectable RR below / above 1 (80% power) |
|---|---|
| P1 canakinumab | 0.59 / 1.69 |
| P2/H5 anti-NGF | 0.72 / 1.40 |
| P3 SGLT2i (gout) | 0.57 / 1.76 |
| H1 GLP-1RA | 0.84 / 1.19 |
| H2 SGLT2i | 0.80 / 1.25 |
| H3 DPP-4i | 0.71 / 1.42 |
| H4 IL-1 inhibition | 0.59 / 1.69 |
| H6 bone-active agents | 0.74 / 1.35 |
| H7 statin + PCSK9i | 0.80 / 1.24 |
| H8 VKA vs DOAC | 0.78 / 1.28 |

#### (b) GLP-1 receptor agonists by molecule (Cochran Q between molecules 11.7, df 8, P = 0.17)

| Molecule | Trials | Events (A vs C) | RR (95% CI) |
|---|---|---|---|
| tirzepatide | 5 | 5 vs 6 | 0.52 (0.17–1.53) |
| albiglutide | 3 | 10 vs 17 | 0.50 (0.22–1.12) |
| dulaglutide | 3 | 137 vs 110 | 1.24 (0.97–1.59) |
| lixisenatide | 3 | 8 vs 8 | 0.75 (0.25–2.24) |
| orforglipron | 3 | 3 vs 2 | 0.80 (0.15–4.35) |
| exenatide | 2 | 70 vs 79 | 0.89 (0.65–1.23) |
| semaglutide | 19 | 173 vs 191 | 0.86 (0.70–1.06) |
| liraglutide | 10 | 98 vs 74 | 1.15 (0.85–1.56) |
| efpeglenatide | 1 | 4 vs 4 | 0.50 (0.12–1.99) |

### Supplementary Table S12. Analyses requested by simulated peer review (post hoc; Protocol addendum D9, defined before they were run)

#### (a) Head-to-head pairs listed in protocol section 4 and not run before (deviation D5)

| Analysis | Trials (informative) | Events (A vs C) | RR (95% CI) | P | From previewed trials |
|---|---|---|---|---|---|
| tirzepatide vs other GLP-1RA (all trials) | 4 (3) | 33 vs 49 | 0.66 (0.43–1.03) | 0.069 | 96% |

| | | | | | |
|---|---|---|---|---|---|
| tirzepatide vs dulaglutide, SURPASS-CVOT (NCT04255433) | 1 (1) | 30 vs 49 | 0.61 (0.39–0.96) | 0.034 | 100% |
| tirzepatide vs other GLP-1RA without SURPASS-CVOT | 3 (2) | 3 vs 0 | — | | 0% |
| DPP-4i vs sulfonylurea (all trials) | 8 (7) | 71 vs 78 | 0.87 (0.63–1.20) | 0.39 | 89% |
| linagliptin vs glimepiride, CAROLINA (NCT01243424) | 1 (1) | 54 vs 66 | 0.81 (0.57–1.16) | 0.26 | 100% |
| DPP-4i vs sulfonylurea without CAROLINA | 7 (6) | 17 vs 12 | 1.15 (0.56–2.37) | 0.71 | 41% |

#### (b) Capture of the outcome: osteoarthritis-related SAEs per 1,000 person-years in control arms (adverse-event window taken as follow-up)

| Measure | Trials | Median (IQR) or ratio |
|---|---|---|
| control-arm rate per 1,000 PY, >= 2,000 controls and window >= 104 wk (median) | 70 | 1.38 (0.80–2.29) |
| control-arm rate per 1,000 PY, window >= 52 wk (median) | 559 | 0.00 (0.00–1.41) |
| control-arm rate per 1,000 PY, Cardiovascular / renal / metabolic | 197 | 1.12 (0.00–2.29) |
| control-arm rate per 1,000 PY, Musculoskeletal / pain | 63 | 0.00 (0.00–2.35) |
| control-arm rate per 1,000 PY, Neuro / psychiatric | 48 | 0.00 (0.00–0.86) |
| control-arm rate per 1,000 PY, Oncology | 149 | 0.00 (0.00–0.26) |
| control-arm rate per 1,000 PY, Other | 65 | 0.00 (0.00–1.65) |
| control-arm rate per 1,000 PY, Respiratory | 37 | 0.00 (0.00–0.97) |
| CANTOS placebo arm: registry events / published replacements | 1 | 0.76 |

For comparison, 631 of 5,125 Look AHEAD participants (12%), adults with type 2 diabetes and overweight, reported a total knee replacement during follow-up through the extension study. CANTOS: registry placebo-arm events against replacements implied by the published placebo rate (0.54 per 100 person-years over a median 3.7 years in 3,344 participants).

#### (c) Pre-specified estimates restricted to trials at or above the median combined-arm rate (all trials with a stated window; as defined in the addendum)

| Analysis | Trials (informative) | Events (A vs C) | RR (95% CI) | P | From previewed trials |
|---|---|---|---|---|---|
| P1 canakinumab | 3 (3) | 59 vs 52 | 0.57 (0.39–0.82) | 0.003 | 98% |
| P2/H5 anti-NGF | 6 (6) | 544 vs 66 | 2.39 (1.86–3.07) | 9.4e-12 | 1% |
| P3 SGLT2i (gout) | 35 (15) | 44 vs 57 | 0.68 (0.45–1.01) | 0.053 | 95% |
| H1 GLP-1RA | 38 (36) | 498 vs 483 | 0.97 (0.86–1.10) | 0.64 | 93% |
| H2 SGLT2i | 35 (33) | 343 vs 280 | 0.96 (0.82–1.12) | 0.60 | 92% |
| H3 DPP-4i | 11 (10) | 90 vs 69 | 1.28 (0.93–1.75) | 0.12 | 89% |
| H4 IL-1 inhibition | 3 (3) | 59 vs 52 | 0.57 (0.39–0.82) | 0.003 | 98% |
| H6 bone-active agents | 8 (8) | 154 vs 160 | 0.95 (0.77–1.19) | 0.67 | 52% |

| | | | | | |
|---|---|---|---|---|---|
| H7 statin + PCSK9i | 23 (23) | 335 vs 294 | 1.09 (0.93–1.27) | 0.31 | 46% |
| H8 VKA vs DOAC | 6 (6) | 44 vs 70 | 1.08 (0.74–1.57) | 0.69 | 61% |
| M1 GLP-1RA -> gallbladder | 38 (33) | 798 vs 516 | 1.35 (1.21–1.51) | 1.7e-07 | 86% |
| M2 SGLT2i -> ketoacidosis | 35 (21) | 164 vs 71 | 1.76 (1.32–2.35) | 1.0e-04 | 69% |
| M3 TZD -> heart failure | 3 (2) | 13 vs 9 | 1.16 (0.48–2.83) | 0.74 | 0% |
| M5 SGLT2i -> heart failure | 35 (27) | 3086 vs 3876 | 0.74 (0.71–0.77) | 5.3e-42 | 95% |

#### (d) As (c), restricted to the upper half of trials with at least one event (added after (c), which mainly removed trials without events)

| Analysis | Trials (informative) | Events (A vs C) | RR (95% CI) | P | From previewed trials |
|---|---|---|---|---|---|
| P1 canakinumab | 2 (2) | 58 vs 52 | 0.56 (0.38–0.81) | 0.002 | 99% |
| P2/H5 anti-NGF | 6 (6) | 544 vs 66 | 2.39 (1.86–3.07) | 9.4e-12 | 1% |
| P3 SGLT2i (gout) | 20 (5) | 12 vs 17 | 0.54 (0.26–1.14) | 0.11 | 83% |
| H1 GLP-1RA | 25 (23) | 400 vs 366 | 1.03 (0.90–1.19) | 0.65 | 93% |
| H2 SGLT2i | 20 (18) | 200 vs 151 | 1.06 (0.86–1.31) | 0.59 | 87% |
| H3 DPP-4i | 6 (6) | 9 vs 7 | 1.11 (0.39–3.15) | 0.84 | 0% |
| H4 IL-1 inhibition | 2 (2) | 58 vs 52 | 0.56 (0.38–0.81) | 0.002 | 99% |
| H6 bone-active agents | 7 (7) | 148 vs 145 | 1.01 (0.81–1.27) | 0.92 | 48% |
| H7 statin + PCSK9i | 16 (16) | 237 vs 212 | 1.05 (0.88–1.27) | 0.58 | 43% |
| H8 VKA vs DOAC | 4 (4) | 14 vs 26 | 0.74 (0.38–1.44) | 0.38 | 0% |
| M1 GLP-1RA -> gallbladder | 25 (20) | 638 vs 436 | 1.32 (1.17–1.49) | 1.0e-05 | 87% |
| M2 SGLT2i -> ketoacidosis | 20 (10) | 81 vs 38 | 1.63 (1.10–2.43) | 0.015 | 67% |
| M3 TZD -> heart failure | 1 (1) | 7 vs 8 | 0.86 (0.31–2.37) | 0.77 | 0% |
| M5 SGLT2i -> heart failure | 20 (14) | 621 vs 737 | 0.75 (0.68–0.83) | 7.5e-08 | 73% |

#### (e) Composition of the hypothesis estimates

| Analysis | Trials (informative) | Events (A vs C) | RR (95% CI) | P | From previewed trials |
|---|---|---|---|---|---|
| H1 GLP-1RA, all | 49 (40) | 508 vs 491 | 0.97 (0.86–1.10) | 0.63 | 93% |
| H1 GLP-1RA, SELECT | 1 (1) | 85 vs 80 | 1.06 (0.78–1.44) | 0.70 | 100% |
| H1 GLP-1RA, obesity trials other than SELECT | 21 (18) | 41 vs 21 | 0.97 (0.57–1.68) | 0.93 | 16% |
| H1 GLP-1RA, obesity trials incl. SELECT | 22 (19) | 126 vs 101 | 1.04 (0.80–1.36) | 0.77 | 77% |
| H1 GLP-1RA, diabetes and other trials | 27 (21) | 382 vs 390 | 0.95 (0.83–1.09) | 0.48 | 98% |
| H6 Denosumab | 8 (8) | 155 vs 149 | 1.03 (0.83–1.29) | 0.79 | 47% |
| H6 Bisphosphonate | 4 (2) | 8 vs 3 | 2.66 (0.69–10.20) | 0.15 | 0% |
| H6 Romosozumab | 1 (1) | 6 vs 15 | 0.40 (0.16–1.03) | 0.057 | 100% |
| H7 PCSK9 inhibitor | 22 (22) | 334 vs 292 | 1.09 (0.93–1.27) | 0.29 | 46% |
| H7 Statin | 4 (3) | 19 vs 20 | 0.95 (0.51–1.78) | 0.87 | 85% |
| P2/H5 anti-NGF, all | 12 (11) | 584 vs 77 | 2.31 (1.82–2.92) | 2.8e-12 | 7% |

| | | | | | |
|---|---|---|---|---|---|
| P2/H5 anti-NGF without FACT LTS (NCT02683239) | 11 (10) | 177 vs 33 | 1.55 (1.07–2.25) | 0.021 | 23% |
| P2/H5 anti-NGF, maximum-term counting | 12 (11) | 349 vs 36 | 3.08 (2.19–4.34) | 1.4e-10 | 12% |
| P2/H5 anti-NGF, arthroplasty terms only | 12 (3) | 138 vs 36 | 1.06 (0.73–1.52) | 0.77 | 0% |
| H8 VKA vs DOAC, all | 21 (17) | 240 vs 311 | 1.10 (0.92–1.30) | 0.29 | 87% |
| H8 VKA vs DOAC, atrial fibrillation trials | 14 (10) | 221 vs 294 | 1.09 (0.92–1.31) | 0.32 | 93% |
| H8 VKA vs DOAC, other (venous thromboembolism and other) | 7 (7) | 19 vs 17 | 1.12 (0.58–2.15) | 0.74 | 0% |
| odanacatib (cathepsin K inhibitor) | 1 (1) | 136 vs 115 | 1.18 (0.92–1.51) | 0.19 | 100% |
| centrally acting anti-obesity drugs (ATC A08AA) | 9 (7) | 244 vs 245 | 0.99 (0.83–1.18) | 0.90 | 0% |

#### (f) Calibration as a bracket: null distributions

| Null | Estimates | mean, SD (log scale) |
|---|---|---|
| primary | 100 | +0.106, 0.236 |
| without corticosteroid x cataract (biological) | 97 | +0.066, 0.152 |
| >= 50 events | 33 | +0.087, 0.267 |
| without known-effect estimates (A3, refitted) | 96 | +0.051, 0.126 |

#### (g) Calibrated risk ratios (95% CI); smallest risk ratio detectable with 80% power under each null

| Analysis | >= 50 events | leave-one-class-out | nominal (no calibration) | primary | without corticosteroid x cataract (biological | without known-effect estimates (A3, refitted |
|---|---|---|---|---|---|---|
| H1 GLP-1RA | 0.89 (0.52–1.52); 0.46 | 0.87 (0.55–1.40); 0.51 | 0.97 (0.86–1.10); 0.84 | 0.87 (0.54–1.41); 0.50 | 0.91 (0.66–1.25); 0.63 | 0.92 (0.70–1.21); 0.67 |
| H2 SGLT2i | 0.88 (0.51–1.52); 0.46 | 0.86 (0.52–1.42); 0.49 | 0.96 (0.82–1.12); 0.80 | 0.86 (0.53–1.40); 0.50 | 0.90 (0.64–1.26); 0.62 | 0.91 (0.68–1.22); 0.66 |
| H3 DPP-4i | 1.15 (0.65–2.06); 0.44 | 1.13 (0.66–1.92); 0.47 | 1.26 (0.99–1.61); 0.71 | 1.13 (0.67–1.91); 0.47 | 1.18 (0.80–1.73); 0.58 | 1.20 (0.85–1.69); 0.61 |
| H4 IL-1 inhibition | 0.52 (0.28–0.99); 0.40 | 0.51 (0.28–0.93); 0.43 | 0.57 (0.39–0.82); 0.59 | 0.51 (0.28–0.93); 0.43 | 0.53 (0.33–0.86); 0.51 | 0.54 (0.35–0.84); 0.53 |
| H6 bone-active agents | 0.92 (0.52–1.62); 0.45 | 0.89 (0.54–1.48); 0.49 | 1.00 (0.81–1.24); 0.74 | 0.90 (0.54–1.50); 0.48 | 0.94 (0.65–1.35); 0.59 | 0.95 (0.69–1.32); 0.63 |
| H7 statin + PCSK9i | 0.99 (0.57–1.71); 0.46 | 0.98 (0.60–1.60); 0.49 | 1.08 (0.93–1.26); 0.80 | 0.97 (0.60–1.58); 0.50 | 1.01 (0.72–1.41); 0.62 | 1.03 (0.77–1.37); 0.66 |
| H8 VKA vs DOAC | 1.00 (0.58–1.74); 0.46 | 0.98 (0.59–1.62); 0.49 | 1.10 (0.92–1.30); 0.78 | 0.99 (0.60–1.61); 0.49 | 1.03 (0.73–1.45); 0.61 | 1.04 (0.77–1.41); 0.65 |
| P2/H5 anti-NGF | 2.11 (1.19–3.75); 0.44 | 2.07 (1.23–3.48); 0.48 | 2.31 (1.82–2.92); 0.72 | 2.07 (1.23–3.48); 0.48 | 2.16 (1.48–3.16); 0.58 | 2.19 (1.56–3.08); 0.61 |

Calibration shifts the log risk ratio by the null mean and adds the null variance. The biological null removes the corticosteroid–cataract pairs only; the refitted null (addendum A3) also removes

GLP-1RA–hernia and pairs with nominal P < 0.001; the leave-one-class-out null removes the class's own negative-control estimates.

### Supplementary figure legends

**Supplementary Figure S1. Trial selection and analysis sets.** Trials without any osteoarthritis-related, gout, cataract or inguinal-hernia SAE in any arm carry no information for Mantel–Haenszel estimates and were not adjudicated. D7, D9 and D10, corrections under the first, second and third addenda (Supplementary Files 4, 6 and 7), made after simulated internal review, simulated peer review and an automated re-audit, respectively (Supplementary Table S5). SAE, serious adverse event; VKA, vitamin-K antagonist; DOAC, direct oral anticoagulant.

**Supplementary Figure S2. Positive controls (values for Figure 2A).** Registry Mantel–Haenszel risk ratios (RR; filled squares) and published randomized estimates (open diamonds; CANTOS[15] and meta-analyses[30–35]); events, active vs control. The upper three controls were pre-specified in the protocol; the lower five were added in the first addendum, with their expected effects fixed before extraction (Supplementary File 4). Anti-NGF antibodies have no published randomized estimate for this outcome. GLP-1RA, glucagon-like peptide-1 receptor agonist; SGLT2, sodium–glucose cotransporter-2; DOAC, direct oral anticoagulant; VKA, vitamin-K antagonist; NGF, nerve growth factor; OA, osteoarthritis.

**Supplementary Figure S3. Pre-specified hypotheses H1–H8 (values for Figure 4A).** Mantel–Haenszel RRs, events (active vs control), empirically calibrated 95% CIs (primary null) and Holm-adjusted P values; arrows give the hypothesized direction. Indented rows give the estimate from the trials outside the preliminary analysis and the share of events they contain. H8 is a head-to-head comparison.

**Supplementary Figure S4. Published observational estimates and randomized estimates for the same drug classes (values for Figure 5).** Observational estimates (diamonds; filled, difference at P < 0.05; grey, not counted because the direction cannot be read from the source) and randomized estimates (squares), the P value for their difference and the class of each pair: differs (P < 0.05), compatible (P ≥ 0.05 and randomized 95% CI upper/lower ratio of at most 2) or uninformative. † Adjusted indirect comparison of two placebo-controlled class estimates (Bucher method). Arrows, intervals beyond the axis. Sources (PMIDs) and estimates not counted: Supplementary Table S6.

**Supplementary Figure S5. GLP-1 receptor agonists and osteoarthritis-related SAEs.** (A) The 12 largest trials and the pooled estimate. (B) Trial risk ratios against the adverse-event reporting window, with the inverse-variance-weighted meta-regression line.

**Supplementary Figure S6. GLP-1 receptor agonists and inguinal-hernia SAEs (exploratory),** by population and molecule, with comparator classes and the cataract negative-control outcome.

**Supplementary Figure S7. Class-level risk ratios for osteoarthritis-related SAEs against risk ratios for the negative-control outcomes** cataract (A) and inguinal hernia (B), for classes with at least 10 osteoarthritis-related SAEs; r, unweighted Pearson correlation of log RRs (inverse-variance-weighted values in the Results).

### Supplementary Figure S1

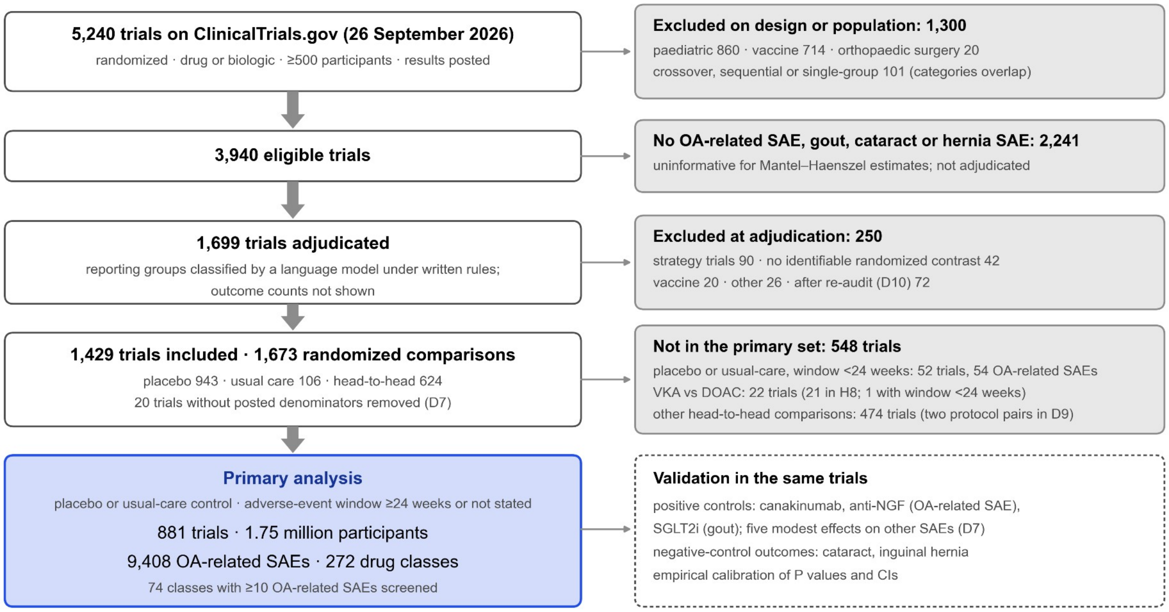

### Supplementary Figure S2

■ registry SAE tables (Mantel–Haenszel) ◇ published randomized estimate

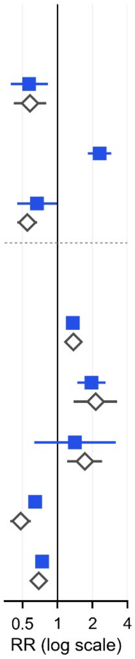

| | Trials | Events | Registry RR | Published |
|---|---|---|---|---|
| **Pre-specified (26 September 2026)** | | | | |
| Canakinumab → OA-related SAE<br>previewed before the protocol | 3 | 59 vs 52 | 0.57 (0.39–0.82) | 0.58 (0.42–0.80) |
| Anti-NGF antibodies → OA-related SAE<br>rapidly progressive OA in 307 of 593 active-arm events | 12 | 584 vs 77 | 2.31 (1.82–2.92) | — |
| SGLT2 inhibitors → gout SAE<br>previewed before the protocol | 46 | 44 vs 58 | 0.66 (0.45–0.99) | 0.55 (0.45–0.67) |
| **Added after review; expected effects fixed before extraction** | | | | |
| GLP-1RA → gallbladder or biliary SAE<br>not an endpoint of the trials | 49 | 862 vs 548 | 1.36 (1.22–1.51) | 1.37 (1.23–1.52) |
| SGLT2 inhibitors → ketoacidosis SAE<br>not an endpoint of the trials | 46 | 188 vs 72 | 1.96 (1.48–2.60) | 2.13 (1.38–3.27) |
| Thiazolidinediones → heart-failure SAE<br>not an endpoint of the trials | 7 | 21 vs 10 | 1.42 (0.63–3.18) | 1.72 (1.21–2.42) |
| DOAC vs VKA → intracranial haemorrhage SAE<br>primary safety endpoint of the trials | 21 | 419 vs 480 | 0.64 (0.56–0.73) | 0.48 (0.39–0.59) |
| SGLT2 inhibitors → heart-failure SAE<br>efficacy endpoint of many trials | 46 | 3,326 vs 4,209 | 0.74 (0.71–0.77) | 0.69 (0.61–0.79) |

### Supplementary Figure S3

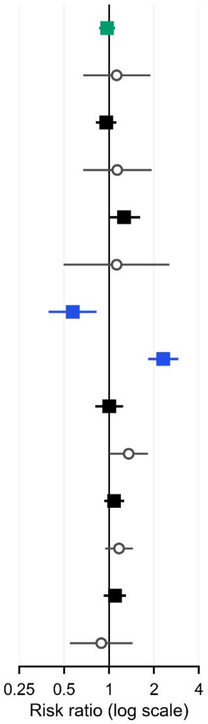

| | Trials | Events | RR (95% CI) | Calibrated | Holm P |
|---|---|---|---|---|---|
| H1 GLP-1 receptor agonists (↓) | 49 | 508 vs 491 | 0.97 (0.86–1.10) | 0.54–1.41 | 1.00 |
| outside the preliminary analysis (7% of events) | 36 | | 1.12 (0.67–1.88) | | |
| H2 SGLT2 inhibitors (two-sided) | 46 | 348 vs 285 | 0.96 (0.82–1.12) | 0.53–1.40 | 1.00 |
| outside the preliminary analysis (10% of events) | 33 | | 1.13 (0.67–1.93) | | |
| H3 DPP-4 inhibitors (↑) | 30 | 150 vs 114 | 1.26 (0.99–1.61) | 0.67–1.91 | 0.38 |
| outside the preliminary analysis (10% of events) | 25 | | 1.12 (0.49–2.53) | | |
| H4 IL-1β inhibition (↓) | 3 | 59 vs 52 | 0.57 (0.39–0.82) | 0.28–0.93 | 0.020 |
| H5 Anti-NGF antibodies (↑) | 12 | 584 vs 77 | 2.31 (1.82–2.92) | 1.23–3.48 | <0.001 |
| H6 Bone-active agents (two-sided) | 13 | 169 vs 167 | 1.00 (0.81–1.24) | 0.54–1.50 | 1.00 |
| outside the preliminary analysis (51% of events) | 11 | | 1.35 (1.00–1.82) | | |
| H7 Statins and PCSK9 inhibitors (two-sided) | 26 | 353 vs 312 | 1.08 (0.93–1.26) | 0.60–1.58 | 1.00 |
| outside the preliminary analysis (51% of events) | 23 | | 1.17 (0.94–1.44) | | |
| H8 Vitamin-K antagonists vs DOACs (↑) | 21 | 240 vs 311 | 1.10 (0.92–1.30) | 0.60–1.61 | 1.00 |
| outside the preliminary analysis (13% of events) | 17 | | 0.89 (0.55–1.44) | | |

### Supplementary Figure S4

◆ observational, differs (P < 0.05) ◇ not counted
◇ observational, no significant difference ■ randomized atlas estimate

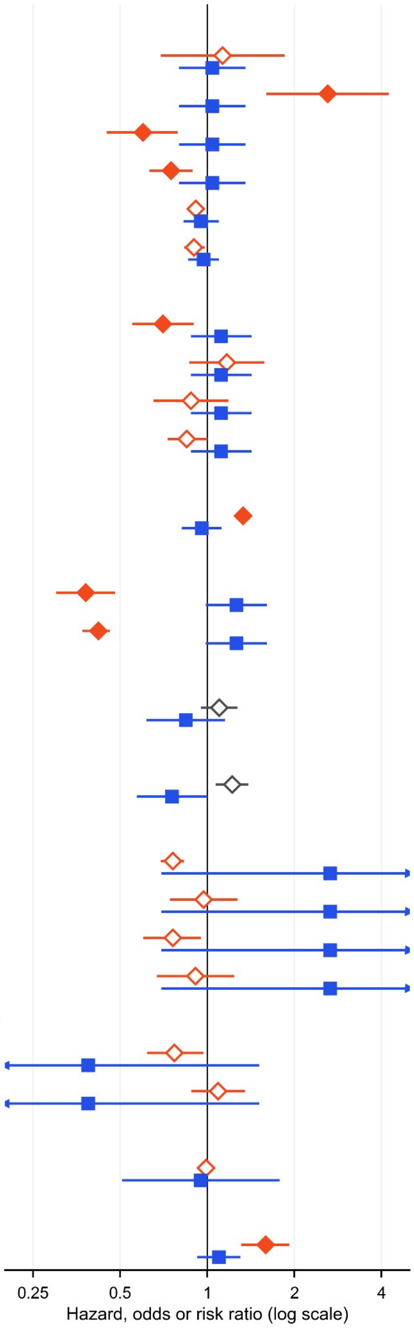

| Study; setting · joint | Observational | Randomized | P | Class |
|---|---|---|---|---|
| **GLP-1RA vs non-use** | | | | |
| Porto 2025<br>obese, no OA · hip | 1.13 (0.69–1.85) | 1.04 (0.80–1.36) | 0.77 | compatible |
| Porto 2025<br>obese, no OA · knee | 2.61 (1.60–4.25) | 1.04 (0.80–1.36) | 0.001 | differs |
| Porto 2025<br>obese, hip OA · hip | 0.60 (0.45–0.79) | 1.04 (0.80–1.36) | 0.005 | differs |
| Porto 2025<br>obese, knee OA · knee | 0.75 (0.63–0.89) | 1.04 (0.80–1.36) | 0.043 | differs |
| Lin 2025<br>T2D, knee OA · knee | 0.91 (0.89–0.98) | 0.95 (0.83–1.09) | 0.59 | compatible |
| Carter 2026<br>knee OA · knee | 0.90 (0.83–0.98) | 0.97 (0.86–1.10) | 0.32 | compatible |
| **SGLT2i vs GLP-1RA** | | | | |
| Su 2025<br>T2D · hip or knee | 0.70 (0.55–0.90) | 1.12 (0.88–1.42) † | 0.008 | differs |
| Jao 2026<br>T2D · hip | 1.17 (0.86–1.58) | 1.12 (0.88–1.42) † | 0.82 | compatible |
| Jao 2026<br>T2D · knee | 0.88 (0.65–1.18) | 1.12 (0.88–1.42) † | 0.22 | compatible |
| Liu 2026<br>T2D · any joint | 0.85 (0.73–1.00) | 1.12 (0.88–1.42) † | 0.063 | compatible |
| **SGLT2i vs non-use** | | | | |
| Hu 2026<br>T2D · knee | 1.33 (1.25–1.42) | 0.96 (0.82–1.12) | <0.001 | differs |
| **DPP-4i vs non-use** | | | | |
| Chen 2026<br>T2D · hip | 0.38 (0.30–0.48) | 1.26 (0.99–1.61) | <0.001 | differs |
| Chen 2026<br>T2D · knee | 0.42 (0.37–0.46) | 1.26 (0.99–1.61) | <0.001 | differs |
| **SGLT2i vs DPP-4i** | | | | |
| Liu 2026<br>T2D · any joint | 1.10 (0.95–1.27) | 0.84 (0.62–1.15) † | 0.13 | not counted |
| **GLP-1RA vs DPP-4i** | | | | |
| Liu 2026<br>T2D · any joint | 1.22 (1.07–1.39) | 0.75 (0.57–1.00) † | 0.002 | not counted |
| **Bisphosphonate vs non-use** | | | | |
| Fu 2017<br>osteoporosis, knee OA · knee | 0.76 (0.69–0.83) | 2.66 (0.69–10.20) | 0.068 | uninformative |
| Hung 2018<br>hip OA · hip | 0.97 (0.74–1.27) | 2.66 (0.69–10.20) | 0.15 | uninformative |
| Neogi 2018<br>women, knee OA · knee | 0.76 (0.60–0.95) | 2.66 (0.69–10.20) | 0.072 | uninformative |
| Wang J 2026<br>UK Biobank · knee | 0.91 (0.67–1.24) | 2.66 (0.69–10.20) | 0.13 | uninformative |
| **Denosumab vs bisphosphonate** | | | | |
| Yu 2024<br>women, knee OA · knee | 0.77 (0.62–0.97) | 0.39 (0.10–1.51) † | 0.33 | uninformative |
| Chen JY 2025<br>women, post-traumatic knee OA · knee | 1.09 (0.88–1.35) | 0.39 (0.10–1.51) † | 0.14 | uninformative |
| **Statin vs non-use** | | | | |
| Sarmanova 2020<br>general population · hip or knee | 0.99 (0.97–1.03) | 0.95 (0.51–1.78) | 0.89 | uninformative |
| **Warfarin vs DOAC** | | | | |
| Ballal 2021<br>atrial fibrillation · hip or knee | 1.59 (1.31–1.92) | 1.10 (0.92–1.30) | 0.005 | differs |

### Supplementary Figure S5

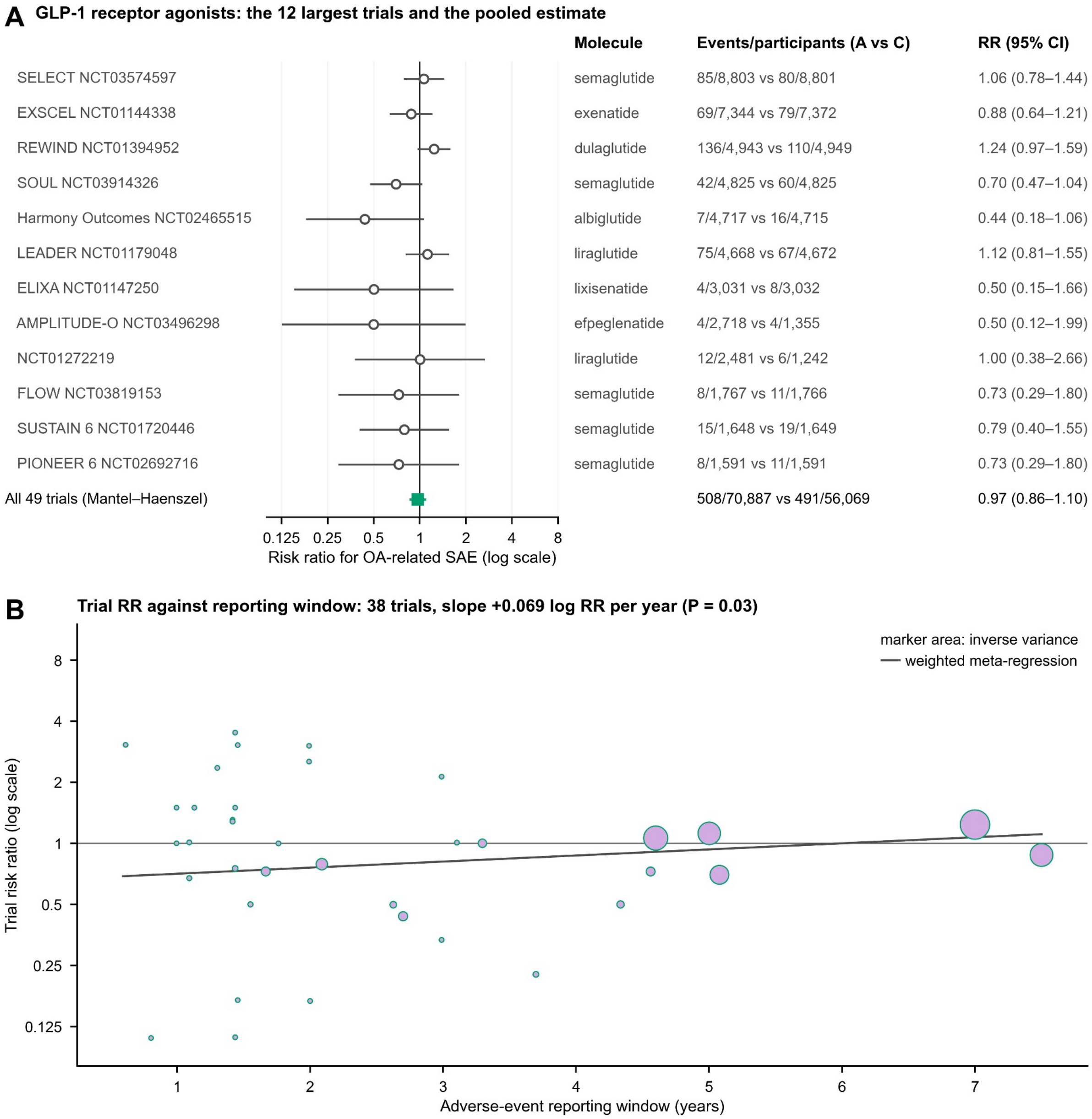

### Supplementary Figure S6

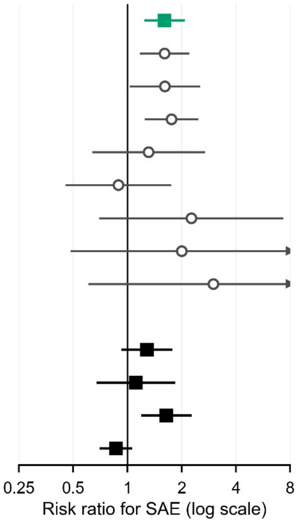

| | Trials | Events (A vs C) | RR (95% CI) | P (nominal) |
|---|---|---|---|---|
| GLP-1RA, all trials | 49 | 152 vs 89 | 1.61 (1.24–2.08) | <0.001 |
| type 2 diabetes and other trials | 27 | 103 vs 61 | 1.61 (1.17–2.20) | 0.003 |
| obesity trials | 22 | 49 vs 28 | 1.61 (1.03–2.53) | 0.039 |
| semaglutide | 16 | 89 vs 49 | 1.75 (1.25–2.47) | |
| dulaglutide | 3 | 17 vs 13 | 1.31 (0.64–2.69) | |
| liraglutide | 9 | 18 vs 17 | 0.89 (0.45–1.75) | |
| exenatide | 2 | 9 vs 4 | 2.26 (0.70–7.33) | |
| lixisenatide | 3 | 7 vs 3 | 2.00 (0.48–8.30) | |
| albiglutide | 3 | 6 vs 2 | 3.00 (0.61–14.85) | |
| **Comparator classes and outcome** | | | | |
| SGLT2 inhibitors, inguinal hernia | 46 | 95 vs 63 | 1.28 (0.93–1.77) | 0.13 |
| DPP-4 inhibitors, inguinal hernia | 30 | 33 vs 29 | 1.11 (0.67–1.84) | 0.67 |
| PCSK9 inhibitors, inguinal hernia | 22 | 101 vs 60 | 1.65 (1.20–2.27) | 0.002 |
| GLP-1RA, cataract (other negative control) | 49 | 168 vs 187 | 0.86 (0.70–1.06) | 0.17 |

### Supplementary Figure S7

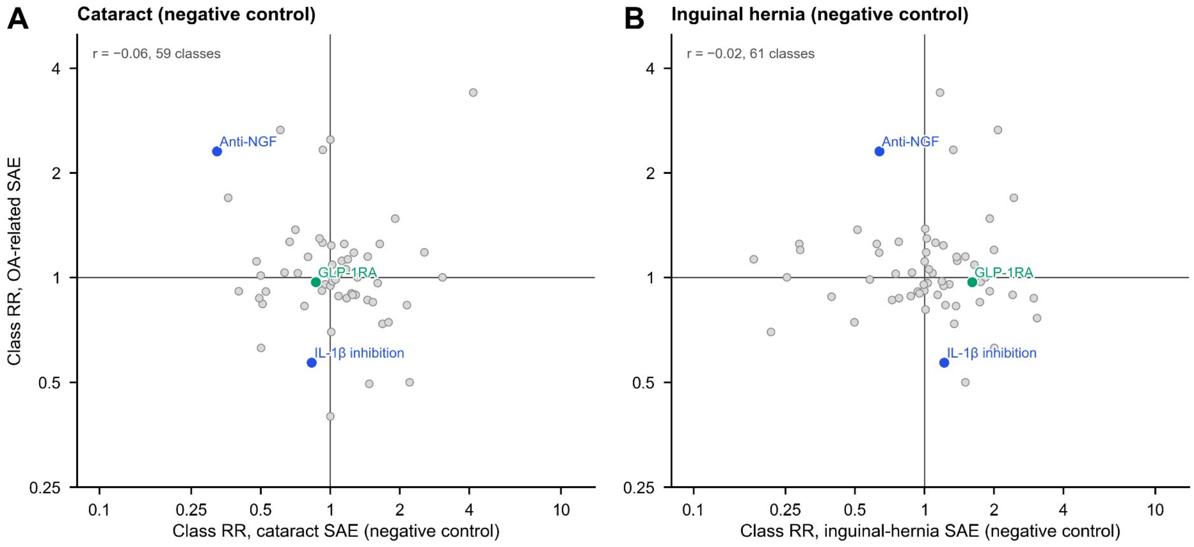
